# The contribution of de novo coding mutations to meningomyelocele

**DOI:** 10.1101/2024.02.28.24303390

**Authors:** Yoo-Jin Ha, Isaac Tang, Ashna Nisal, Ishani Jhamb, Cassidy Wallace, Sarah Schroeder, Chanjae Lee, Keng loi Vong, Naomi Meave, Fiza Jiwani, Chelsea Barrows, Sangmoon Lee, Nan Jiang, Arzoo Patel, Francisco A. Blanco, Seyoung Yu, Hui Su Jeong, Isaac Plutzer, Michael B. Major, Béatrice Benoit, Christian Poüs, Caleb Heffner, Zoha Kibar, Gyang Markus Bot, Hope Northrup, Kit Sing Au, Madison Strain, Allison Ashley-Koch, Richard H. Finnell, Joan T. Le, Hal Meltzer, Camila Araujo, Helio R. Machado, Roger E. Stevenson, Anna Yurrita, Sara Mumtaz, Osvaldo M. Mutchinick, José Ramón Medina-Bereciartu, Friedhelm Hildebrandt, Gia Melikishvili, Rony Marwan, Valeria Capra, Mahmoud M. Noureldeen, Aida M.S. Salem, Mahmoud Y. Issa, Maha S. Zaki, Ji Eun Lee, Anna Alkelai, Alan R. Shuldiner, Stephen F. Kingsmore, Stephen A. Murray, Heon Yung Gee, W. Todd Miller, Kimberley F. Tolias, John B. Wallingford, Spina Bifida Sequencing Consortium, Sangwoo Kim, Joseph G. Gleeson

## Abstract

Meningomyelocele (MM) is considered a genetically complex disease resulting from failure of neural tube closure (NTD). Patients display neuromotor disability and frequent hydrocephalus requiring ventricular shunting. A few proposed genes contribute to disease susceptibility, but most risk remains unexplained^1^. We postulated that de novo mutations (DNMs) under purifying selection contribute to MM risk^2^. Here we recruited a cohort of 851 MM trios requiring shunting at birth, compared with 732 control trios, and found that de novo likely gene disrupting or damaging missense mutations occur in approximately 22.3% of subjects, 28% of which are estimated to contribute to disease risk. The 187 genes with damaging DNMs collectively define networks including actin cytoskeleton and microtubule-based processes, axon guidance, and histone modification. Gene validation demonstrates partial or complete loss of function, impaired signaling and defective neural tube closure in *Xenopus* embryos. Our results suggest DNMs make key contributions to MM risk, and highlight critical pathways required for neural tube closure in human embryogenesis.

## Main

MM, also known as spina bifida, is the most common central nervous system (CNS) structural defect in humans. MM results from failed closure of the neural tube in the first six weeks of gestation. While folic acid supplementation has reduced disease burden^3^, the incidence of 1:3000–10,000 live-births, along with the associated lifelong neuromotor disability and increased mortality, has focused attention on this condition. Timely diagnosis allows for prenatal counseling and informed management choices, including termination of pregnancy, fetal surgery, or postnatal surgery for the nearly fully concordant hydrocephalus accompanying the hindbrain Chiari type II malformation^4,5^.

Over 20 million people worldwide live with an NTD yet causes remain poorly defined. Mouse knockout studies established hundreds of genes associated with NTD, often with partial penetrance, implicating apical-basal polarity, Wnt/PCP, Wnt/beta-catenin, and DNA transcription pathways^6^. Despite epidemiological heritability estimates of 60-70% for NTDs^7^, few genes are implicated in human NTDs, potentially due to genetic heterogeneity. Additive effects of common SNPs have been proposed, especially in the folate metabolism gene *MTHFR* (C677T, p.Ala222Val), but meta-analysis from 22 different association studies suggest only a modest risk (OR 1.23, 95% CI: 1.07–1.42)^8^. Inherited mutations in *VANGL1/2, TBXT, CCL2, CELSR1* were identified in cohort studies^9–12^, but represent only a few percent of cases. Prior successes in de novo mutation (DNM) approaches in conditions like autism and congenital heart disease, where mutations are potentially under purifying selection^13,14^, prompted a trio approach in 43 families, identifying recurrent mutations in *SHROOM3*^15^, but left unanswered whether larger studies would be better powered.

### Establishing the Spina Bifida Sequencing Consortium

A critical step in DNM discovery is assessment for an excess burden of damaging mutations in cases vs. controls^13^. With the conservative assumptions of a modest increase in damaging DNMs in cases and estimating perhaps 50–100 total MM genetic risk loci, we calculated that a cohort of 300–500 trios would be required to identify 5–10 recurrently mutated genes (see **Supplementary Information** and **Extended Data Fig. 1**). In 2015, we thus established the Spina Bifida Sequencing Consortium (SBSC) to aggregate prior CDC (Centers for Disease Control and Prevention, USA)-supported trio recruitment efforts, alongside targeted new recruitment from specialists in more than 12 countries, and through social media outreach. Recruitment was limited to diagnosis of MM probands with open neural tissue observed at birth and hydrocephalus requiring shunting, specifying the most severe form of MM compatible with long term survival (**Methods**). Our ethnically diverse cohort was designed specifically to assess risk from DNMs using trio sequencing, matched with 732 control trios from the Simons Simplex Collection^16^. After five years of recruitment, we had enrolled 325 trios, performed trio whole exome sequencing (WES) and DNM analysis, but found no recurrently mutated genes, so we extended the recruitment timeline for an additional five years, to double the cohort size.

After 10 years of recruitment, we assembled a cohort of 851 MM trios (839 trios and 6 quartets) consisting of exome sequence data of 2,541 individuals. SBSC was populated by 15 different worldwide sites, incorporating both historic CDC-assembled cohorts and an aggregated earlier report of 39 NTD trios (35 trios and 2 quartets)^15^.

### Excessive damaging DNM burden in MM trios

After a series of strict quality controls and kinship analyses applied to trio exomes, we constructed a high-confidence call set of 2,592 DNMs (1,458 from 777 MM trios, and 1,134 from 725 control trios) (**Methods**), and confirmed strong DNM signatures^17^ (**Extended Data Fig. 2a**–2b). The rates of the total and functionally categorized DNMs (LGD: likely gene disrupting, D-Mis: damaging missense, tolerant missense, and synonymous) were calculated for the jointly covered consensus regions (**Fig. 1a)**, wherein total DNM rates lie in a Poisson distribution (**Extended Data Fig. 2c**–2d, **Methods**). The average mutation rates of total DNMs were 1.22 × 10^-8^ (1.13 × 10^-8^–1.31 × 10^-8^, 95% CI) in MM and 1.18 × 10^-8^ (1.09 × 10^-8^–1.28 × 10^-8^, 95% CI) in controls, which did not differ significantly (**Extended Data Fig. 2e**, Wilcoxon rank-sum test *P* value: 0.56), and were also comparable to the rates from previous studies (1.08 × 10^-8^– 1.32 × 10^-8^)^18,19^.

**Figure 1.**
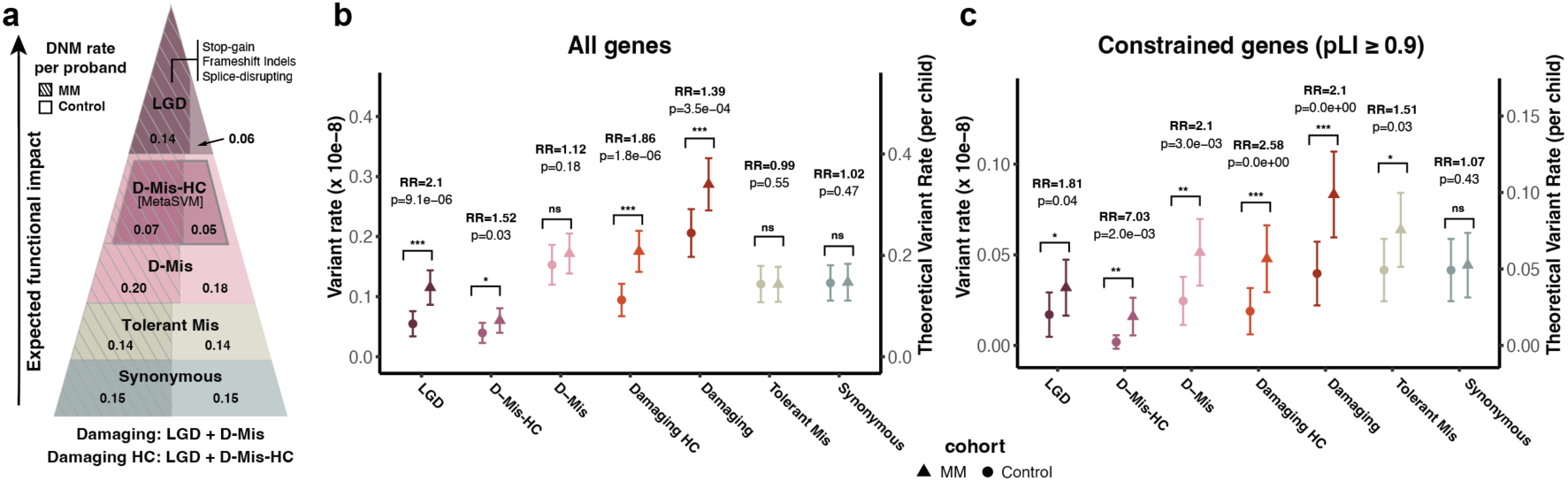
Enrichment of damaging DNMs in MM versus control. **a,** DNMs categorized by predicted functional impact: LGD (likely gene disrupting), D-Mis (damaging missense), D-Mis-HC (damaging missense of high confidence, called from a meta predictor), Tolerant Mis (missense of tolerant functional impact), and Synonymous. DNM rates per proband in MM (left sides of the triangle with strips) and control (right sides). Areas represent the ratio of the DNM rates between MM and controls within each functional category. Hashed: MM; Unhashed: controls. **b–c,** Variant rate (10^-8^) and theoretical rate (DNM rate per child) on left and right y-axis, respectively. Statistical analysis of the ratio of DNM rates between MM (triangles) and controls (circles), denoted by ratio of the two Poisson rates, or rate ratio (RR), calculated within (**b**) ‘All genes’ (*n* = 19,658) and (**c**) ‘Constrained genes’ (*n* = 3,060; pLI ≥ 0.9). Theoretical rate was calculated by normalization of the variant rate with the total size of hg38 coding region (59,281,518 bp). Observed and theoretical DNM rates are marked in the left and right y-axis, respectively. *P* values were calculated by a one-sided rate-ratio test. *P* values: *** < 0.001, ** < 0.01, * < 0.05. ns: not significant.

In contrast, we observed an excessive rate of LGD DNMs (frameshift Indels, stop-gained, and splice donor/acceptor mutations) in MM (1.2 × 10^-9^) compared to controls (5.0 × 10^-10^) (**Fig. 1b**), with a ratio of the two Poisson rates (or rate ratio)^20^ of 2.10 (*P* value: 9.0 × 10^-6^). Case-control comparison of DNM rate per proband (i.e. theoretical rate: 0.12 in MM vs. 0.05 in controls, see **Methods**), suggested that 7.2% (95% CI: 2.96%–11.36%) of MM probands display LGD DNM implicated in MM risk (**Table 1**). Based upon the excessive burden, approximately 52.43% of LGD DNMs in MM probands was estimated to contribute to MM risk (**Methods**), comparable with rates in other complex diseases such as Tourette (∼51.3%) and autism spectrum disorders (∼42%)^13,21^. When LGD DNMs were considered alongside D-Mis DNMs (i.e. LGD + D-Mis; referred to as ‘damaging DNMs’), 28.26% (95% CI: 7.75%–48.76%) were linked to MM risk with excessive burden. Of note, a subset of high confidence D-Mis DNMs (referred to as ‘D-Mis-HC’) based upon pathogenicity prediction, conferred approximately 80% of the total LGD risk burden (rate ratio: 1.86, *P* value: 2.0 × 10^-6^). Paternal and maternal ages, expected to be correlated with DNM occurrence^22,23^, were not correlated with the excessive DNMs burden (**Extended Data Fig. 3a**). Moreover, there was no enrichment in tolerant and synonymous missense DNMs.

**Table 1.** Contribution of DNMs to MM risk. The DNM theoretical rate (i.e., DNM rate per child) of MM (*n* = 772) and control (*n* = 724) trios are shown with the 95% confidence interval (CI) of the rate using a one-sample t-test. The % of cases with DNM mediating risk were calculated by the difference between the theoretical rate of the MM and the control with 95% CI using a two-sample t-test (two-sided). The % of cases with DNM mediating risk was estimated by dividing the difference of theoretical rate between MM and control, by the theoretical rate of MM trios, with the 95% CI using two-sample t-test. The theoretical rates were calculated based on consensus regions for proper comparison of DNM rates. Damaging DNM includes LGD (Likely Gene Disrupting) and D-Mis DNM. Damaging-HC (high confidence) includes LGD and D-Mis-HC (high confidence).

| Variant type | Theoretical rate per child<br>(±95% CI) |  | % of cases with DNM<br>mediating risk (±95% CI) | % of DNM carrying MM<br>risk (±95% CI) |
| --- | --- | --- | --- | --- |
|  | MM (n=772) | Control (n=724) |  |  |
| Likely Gene Disrupting (LGD) | 0.12 (0.09-0.14) | 0.05 (0.03-0.08) | 7.16% (2.96%-11.36%) | 52.43% (21.67%-83.18%) |
| Damaging HC | 0.18 (0.14-0.21) | 0.09 (0.07-0.12) | 9.60% (4.45%-14.75%) | 46.15% (21.38-70.92) |
| Damaging | 0.29 (0.24-0.33) | 0.21 (0.17-0.25) | 9.62.% (2.64%-16.59%) | 28.26% (7.75-48.76) |

In 3,060 genes considered to be highly constrained (i.e. the probability of loss-of-function intolerance: pLI ≥ 0.9), the burden of D-Mis DNMs appeared even more significant (rate ratio: 2.10, *P* = 0.003) (**Fig. 1c**). Furthermore, we observed a substantially increased burden of D-Mis-HC DNMs (rate ratio: 7.7, *P* = 0.002). This contrasts with the DNM contributions of other complex diseases, where LGDs play outsized roles compared with D-Mis DNMs^13^.

To assess whether there is major missed heritability using WES compared with whole genome sequencing (WGS), we recruited an additional 101 trios and 1 quartet, and performed WGS using standard discovery pipelines (**Methods**). We identified non-recurrent de novo SNV/Indels at expected rates of 1.72 × 10^-8^ (vs. 1.2 × 10^-8^–2.4 × 10^-8^ in previous publications^19,24^ ,see **Extended Data Table 1**), encompassing 9 putative splice-disrupting and 41 in putative enhancer/promoter regions (**Supplementary Data**). Copy number variant (CNV) analysis revealed two nonrecurrent deletions and two duplications (> 100kb), a rate comparable to other complex diseases^25,26^ (**Extended Data Fig. 3b** and **Extended Data Table 2**). De novo inversions or translocations were not observed. In contrast, a high LGD rate (0.13 per proband) in coding region was replicated in WGS, with three overlapping genes observed in the WES cohort (*KDM1A*, *ITPR3*, and *GRHL2*). Overall, WGS-based analysis mirrored the findings from WES and did not identify an additional major class of mutation (see **Supplementary Information**).

### Functional convergence of DNM genes in MM

Out of 198 damaging DNMs (82 LGD and 116 D-Mis) found in MM trios, Sanger sequencing was conducted in 86% (170 out of 198) where sufficient DNA was available, yielding 96.5% validation rate, which removed six false DNMs. The remaining 192 damaging DNMs (79 LGD and 113 D-Mis) occurred in 187 unique genes, with only five genes mutated in two separate trios (referred to as ‘doubleton’; *PAX3*, *IRS1*, *ZSWIM6*, *BRSK2*, and *VWA8*) (**Fig. 2a** and **Supplementary Data**). Notably, only *PAX3* was previously implicated in human NTDs^15,27^. Among genes identified as singletons, only *TBXT* (splice donor) and *CELSR1* (frameshift) were previously implicated as inherited NTD risk factors^9,11^. The lack of additional recurrently mutated genes underscores a significant gap in our current understanding of MM genetic risk factors.

**Figure 2.**
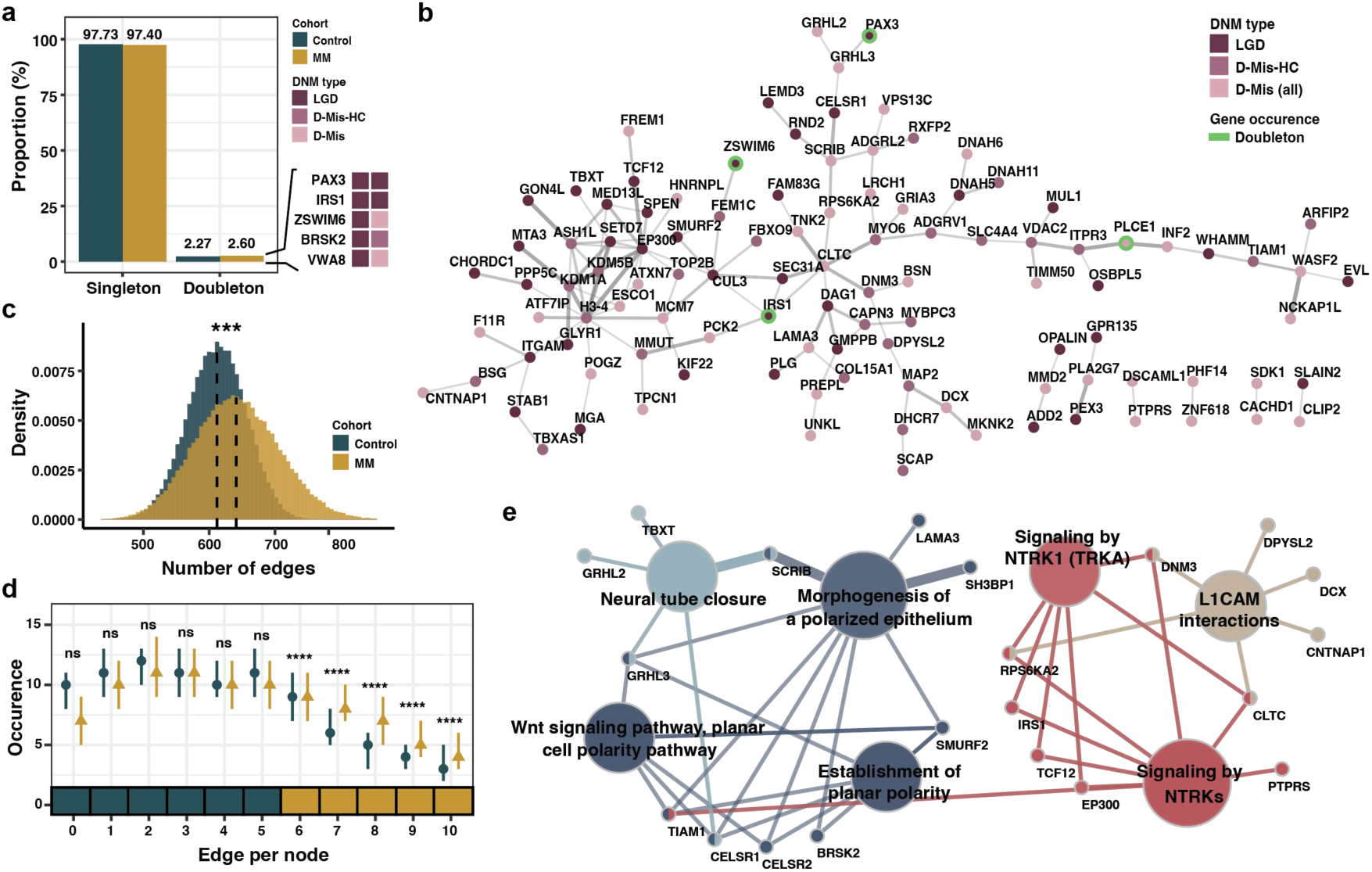
Functional convergence of genes implicated by damaging DNMs. **a,** Proportion of singletons (DNM occurred in one trio) and doubletons (DNMs occurred in two independent trios) damaging (LGD or D-Mis) genes in controls (green) and MM (yellow). Six doubleton genes were annotated with variant functional categories (LGD, D-Mis-HC, or D-Mis). **b,** A protein-protein interaction network composed of MM DNM genes. Genes connected by at least one edge are shown; unconnected (orphan) genes are shown in **Extended Data Fig. 3a**. Node color: variant functional categories: LGD (purple), D-Mis (light pink), and D-Mis-HC (dark pink). Doubleton genes: green borders. Edge thickness: confidence score of the protein interaction, defined in the STRING database. **c**, Distribution of the total number of edges in 100,000 networks randomly generated with bootstrapping (selecting gene set size of 108 (i.e. 80% of the 135 control DNM genes) in control (green) and MM (yellow). Higher number of edges denotes denser network interconnection. *P* values calculated by two-sided Wilcoxon rank-sum test. **d,** Number of nodes with *n* (0 to 10) edges in controls (green) and MM (yellow) gene sets. Error bars: first and third quartile. *P* values calculated by a one-sided Wilcoxon rank-sum test. **e,** GO (Gene Ontology) term network visualization for genes overlapping with 187 MM DNM genes with statistical significance (a.k.a., GO enrichment analysis). GO terms of functional relevance connected by an edge via commonly involved genes (small circles). Node sizes: significance of the terms. Degree of connectivity between terms (edges): kappa statistics by ClueGO. \*\*\*\**P* value < 0.0001. ns: not significant.

Spatial transcriptomic analysis of 36 of the damaging DNM genes confirmed expression at mouse embryonic day 10.5 coinciding with neural tube closure (MERFISH, see **Methods**, **Extended Data Fig. 4**). We found that most genes were expressed broadly in the embryo (86%, 31 out of 36), but a minority (14%) showed some cell type specific expression such as *Celsr1* in neural progenitors and *Stab1* in neural crest progenitors (**Extended Data Fig. 5**, **Supplementary Information**).

We next studied the protein interactions of MM DNM genes using the STRING database^28^ (**Fig. 2b**). Among the 187 genes, 107 (57%) including 4 doubletons, 33 LGDs, and 74 D-Mis were highly interconnected, compared to controls (38% interconnected) (**Extended Data Fig. 6**). Bootstrap analysis confirmed a significantly greater degree of network colocalization than genes in the control group (**Fig. 2c, Methods**, two-sided Wilcoxon rank-sum test, *P* value < 1.0 × 10^-16^). Genes exhibiting a higher node degree (*n* ≥ 6) were predominantly observed in the MM gene set (Wilcoxon rank-sum test, one-sided) (**Fig. 2d**). Indeed, MM DNM genes were significantly enriched in biological pathways such as morphogenesis of polarized epithelium, neuronal cell adhesion, neural tube closure, and signal transduction (Gene Ontology biological processes, adjusted *P* values < 0.05) (**Fig. 2e and Extended Data Table 3**), suggesting functional convergence in neural tube closure.

### DNMs implicate functional modules in neural tube closure

Using the 187 MM DNM genes as seeds, we employed network propagation to construct a comprehensive gene network associated with human MM risk (**Extended Data Fig. 7**, see **Methods**)^29^, by implicating functionally related genes that might not be directly observable in patients due to, for instance, lethal mutations. The propagated network ‘Meningomyel-ome’ comprised 439 genes, including 257 propagated genes, and exhibited 2,447 interconnected edges. We found an over-representation of 374 experimentally proven mouse NTD causal genes^30^ (**Supplementary Data**) in both seed and propagated genes (11 and 17 of the mouse NTD genes in before- and after propagation, one-tailed hypergeometric *P* value: 0.0015), corroborating functional relatedness to human MM. In contrast, no such relatedness was found with propagation in genes carrying DNMs in control trios (*P* value: 0.295).

Applying a clustering algorithm^31^ to the ‘Meningomyel-ome’, we identified five subnetworks in which damaging DNMs and propagated genes were closely connected and enriched within predefined signaling pathways (referred to as submodules): RHO/RAC1 GTPase based actin cytoskeleton organization, microtubule-based process, neuronal migration and axon guidance, metabolism of lipids, and histone modification (FDR < 10^-5^, **Fig. 3a**, see **Methods**)^6,32–38^, suggesting that disrupting these pathways can increase MM risk. Notably, a higher haploinsufficiency of the 51 propagated genes within the submodules (pLI = 0.98 vs. 0.69 of 46 seed genes in median, see **Extended Data Fig. 8**) substantiated the utility of network propagation to further implicate potentially lethal MM risk genes.

**Figure 3.**
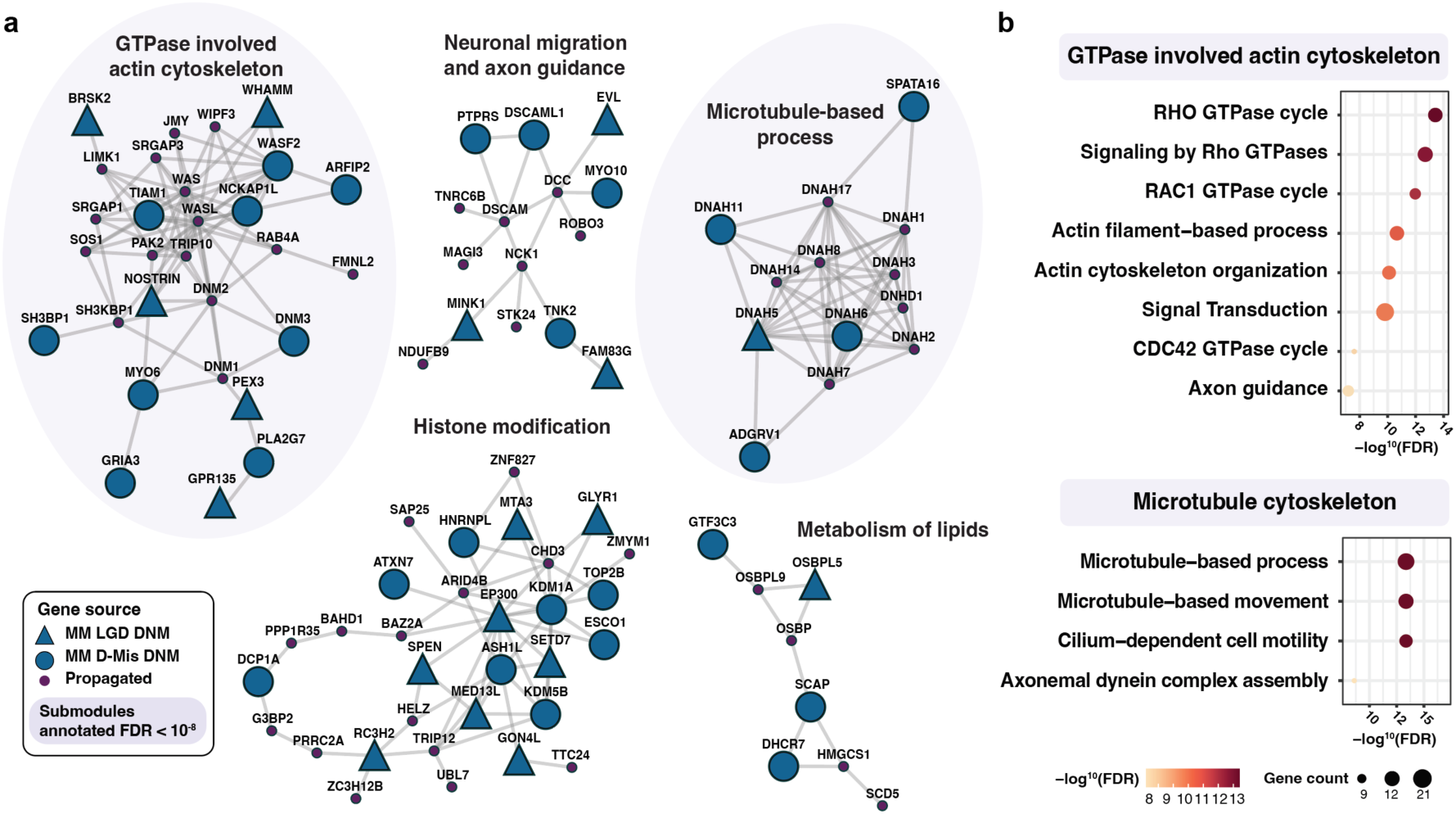
Functional modules that contribute to MM risk. **a,** Functional submodules clustered from the propagated network, ‘Meningomyel-ome’, with the 187 damaging DNM genes from MM cohort used as seeds. Clustered with a Leiden algorithm with co-expression value from STRING database as attributes, five submodules annotated FDR < 10^-6^ by Gene Ontology (GO) Biological Process, KEGG, or Reactome databases shown. **b,** Topmost significantly enriched GO Biological Processes with FDR < 10^-8^ shown. FDR value: -log^10^ scale. LGD DNMs genes: blue triangle. D-Mis DNMs: blue circles. Propagated genes: small purple circles. Functional terms annotated with FDR < 10^-8^ colored with light purple background.

Common among the twelve most strongly associated signaling pathways with FDR below 10^-8^ was actin and microtubules organization processes (**Fig. 3b**). Functional relatedness to other submodules of higher-level processes^39,40^ and essential roles in the neural fold adhesion and closure^41,42^ suggest that RHO/RAC1 GTPase-mediated actin and microtubule organization processes contribute to human MM risk.

### Functional validation of candidate MM genes

We next performed functional assessment of candidate MM DNM alleles or genes. Seven genes—*TNK2*, *TIAM1*, *PLCE1, BRSK2*, *CLIP2, VWA8,* and *WHAMM*—were selected by their relevance to the RHOA/RAC1/CDC42-mediated actin and microtubule polymerization, five of which are direct regulators (**Fig. 4a, Methods**). Patient mutations in these genes were all D-Mis DNMs usually in regulatory domains predicted to interfere with protein function, except for LGD mutation in *WHAMM* predicted to truncate the protein. TNK2 is a CDC42-activated kinase that phosphorylates WASP among other targets. We found that immunoprecipitated TNK2 p.P186L DNM had approximately 60% reduced kinase activity to a WASP pseudo substrate, as benchmarked against wildtype and kinase-dead versions (**Fig. 4b** and **Extended Data Fig. 9**). TIAM1 is a Src-activated RAC1-specific guanine nucleotide exchange factor that promotes filamentous actin (F-actin) assembly. We found that wild type TIAM1 but not the TIAM1 p.H1149P DNM co-localized with filamentous actin (**Fig. 4c-d**). While basal Rac1 activation was unperturbed, p.H1149P showed significant reduction in Rac1 activation following co-expression with constitutively activate Src (**Fig. 4e** and **Extended Data Fig. 10)**. VWA8 is a recurrently mutated mitochondrial matrix-targeting ATPase. We found that the p.R230G proband mutation failed to produce VWA8 protein evidenced on Western blot when transfected in HEK293T cells (**Extended Data Fig. 11**). Assessment of CLIP2, BRSK2 and PLCE1 MM proband mutations did not report robust defective protein function in standard assays (**Extended Data Fig. 12**-14). Interestingly the DNM mutations showing impaired protein function have previously been directly or indirectly linked to NTDs in animal models (see **Supplementary Information**). Thus, half of the six missense mutations showed clear disrupted function, consistent with our predicted rate of DNMs contributing to MM risk (52.43% of LGD, and 28.26% of damaging DNMs).

**Figure 4.**
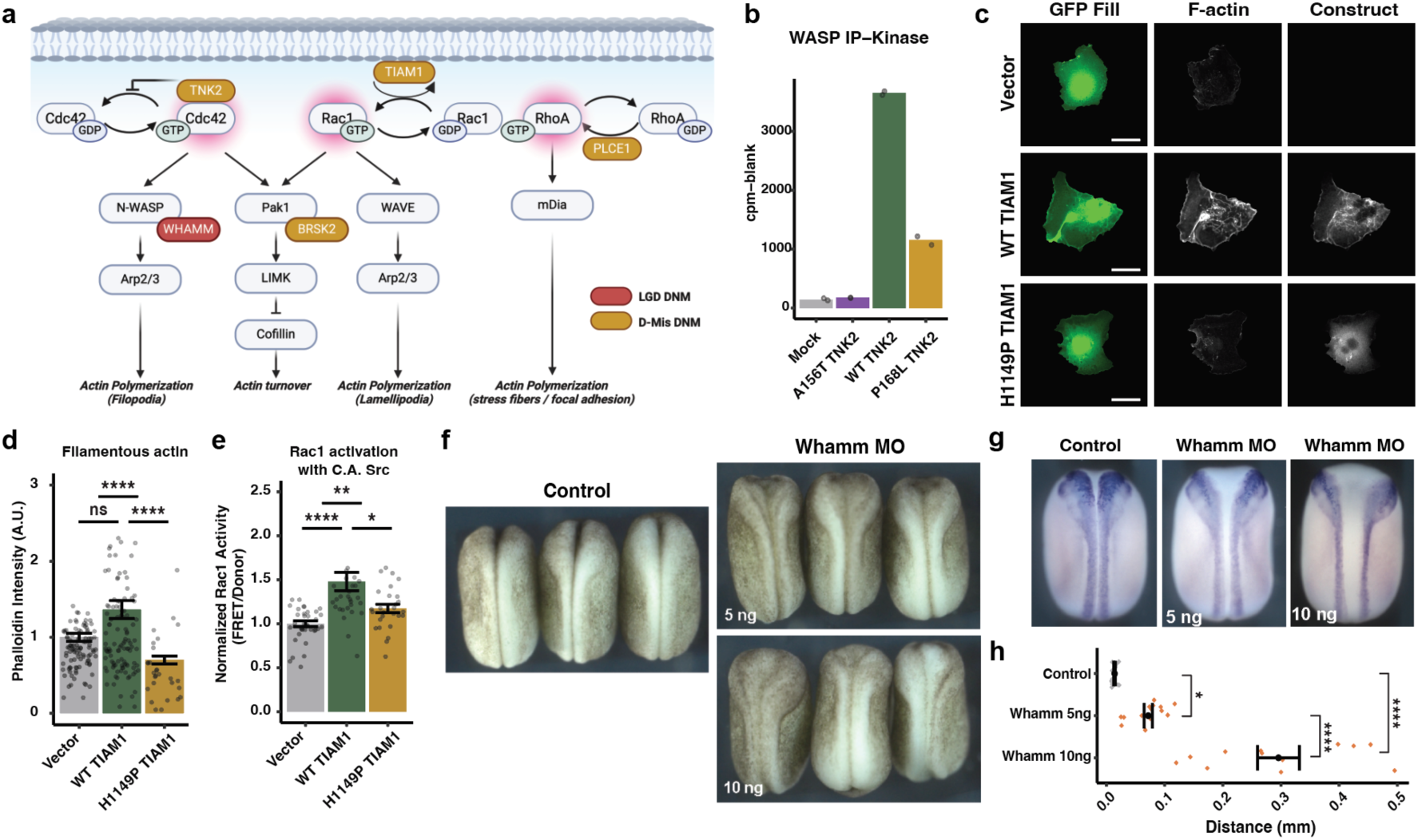
Functional validation of damaging DNMs related to actin polymerization. **a**, Damaging DNM genes highlight ‘GTPase involved actin cytoskeleton pathway’ mediated by WASP and WAVE, involving three GTPases (CDC42, RAC1, and RHOA) and five mutated genes *TNK2, TIAM1, WHAMM, PLCE1, and BRSK2*. Red circle: LGD DNM, Dark yellow circle: D-Mis DNM. **b**, TNK2 p.P168L patient mutation impaired WASP phosphorylation from immunoprecipitation (IP) kinase assay, compared to WT and kinase dead p.A156T. **c–d**, TIAM1 p.H1149P patient mutation showed fewer lamellipodia (F-actin) compared to WT, (**c**) imaged with Phalloidin and (**d**) quantified. Kruskal-Wallis followed by a pairwise Wilcoxon test, *P* value adjusted with Bonferroni. Data shown with Hampel filter and images masked to remove other cells. **e**, TIAM1 p.H1149P patient mutation decreased Rac1 activation with constitutively active (C.A.) Src, observed with Förster resonance energy transfer (FRET). Kruskal-Wallis followed by a pairwise Wilcoxon test, *P* value adjusted with Bonferroni. Data shown with Hampel filter. **f**, Dorsal views of *Xenopus laevis* embryos injected with Whamm morpholino (MO) at Stage 19. Neural tube closure phenotypes observed in a dose-dependent manner with 5 and 10 ng MO. **g**, Neural folds visualized by *pax3* in situ hybridization in the late neural stage. **h,** Quantification of distance between neural folds. Newman-Keuls (two-sided) for control versus multiple conditions after ANOVA. *P* values: **** < 0.0001, *** < 0.001, ** < 0.01, * < 0.05. ns: not significant, LGD: likely-gene disrupting, D-Mis: damaging missense; WT: wild-type.

An LGD mutation found in *WHAMM* was selected for *in vivo* functional assessment in a *Xenopus laevis* neural tube closure model. Splice-blocking morpholino (MO) of the orthologous *Whamm* protein (**Methods**) resulted in dose-dependent NTDs (**Fig. 4f-h**). Specifically, injection of 5 ng of MO led to mild NTD, while 10 ng injection resulted in more severe open NTD in both anterior and posterior regions, as evidenced by *pax3* in situ hybridization, and was rescued by expression of wildtype *Whamm* (**Extended Data Fig. 15)**. These findings confirm the role of at least one of the DNM genes in MM pathogenesis *in vivo* and underscore potential gene dosage effects contributing to MM risk.

### Expanding clinical phenotypes to include MM

We noted that 82 of the MM DNM genes were previously implicated in syndromic or nonsyndromic phenotypes in the Online Mendelian Inheritance in Man (OMIM) database ^43^, raising the question of whether our MM subjects might also express some of these OMIM phenotypes. By recontacting families, we found that 6% (5 out of 83) of the MM subjects (i.e. with mutations in *ZSWIM6*, *PAX3* in two subjects, *TCF12* and *BICRA*) had clinical features in addition to MM (Acromelic frontonasal dysostosis, Waardenburg, Craniosynostosis and Coffin-Siris syndromes, respectively), that suggest MM as a phenotyping expansion (**Supplementary Table 1**). Some subjects were lost to follow up, leading to 21.6% of the phenotypes that could be excluded due to lack of full clinical information. An additional 2 subjects (2.4%) shared some clinical features with OMIM phenotypes but insufficient to make the clinical diagnoses. For the majority of these 82 subjects, the zygosity did not match the OMIM zygosity (i.e. dominant vs. recessive), so it was not surprising that 70% of subjects lacked any OMIM clinical features. We conclude that MM can infrequently present as partially penetrant phenotype in established OMIM disorders, and suggest that specific alleles or environmental factors may determine expressivity.

## Discussion

In this study, we present the first large-scale assessment of DNMs contributing to MM risk. Approximately 22% of MM probands had damaging DNMs, of which about 28% were expected to contribute to MM risk. This risk is comparable to other severe pediatric conditions that are likely under strong purifying selection, as the mutations are unlikely to propagate to offspring^44–46^. In addition to the LGD DNMs found in other severe pediatric conditions, we found strong enrichment of D-Mis DNMs, suggesting that some LGD mutations in these genes may be lethal during embryogenesis. It would be interesting to assess the contribution of D-Mis mutations in other severe pediatric conditions.

Although only a subset of the genes identified likely contribute risk, we found that candidate MM genes were more interconnected than expected by chance. This lends support to the existence of a ‘Meningomyel-ome’, characterized by numerous genes that have potential to affect phenotype, interconnected by regulatory or protein-interaction networks, aligning with an omnigenic model^47^. Many of the networks highlight pathways previously implicated from mouse or frog models, including Wnt signaling, planar cell polarity, actin regulation and cell signaling. Notably, there was little overlap of mutated genes in humans with animal NTD models. One difference is that mouse and frog NTD genes are often identified as lethal embryonic phenotypes^48,49^, whereas our ascertainment was limited to living subjects. Thus, while the genes implicated in laboratory animals vs. human NTDs may be different, we expect the pathways to be highly similar, evident in the Meningomyel-ome. Future work could introduce human DNMs into vertebrate models or human neural tube stem cell models, to bridge this divide.

Of the 187 genes with DNMs in MM subjects, 83 were previously implicated in human phenotypes according to OMIM. Only two of these OMIM conditions listed MM as part of the associated spectrum, and in only five subjects was the overlap sufficient for clinical diagnosis of the linked OMIM phenotype. This suggests that MM should be considered as part of the clinical continuum of a variety of OMIM conditions, which may be expressed in an allelic- or zygosity-specific fashion. Surprisingly, only five of the 187 genes were recurrently mutated in our cohort, suggesting there could be thousands of genes contributing to MM risk (**Supplementary Information**).

Our findings do not resolve the pathogenesis of most MM cases. Rare and de novo copy-number variants^50^, or inherited variants could also contribute to risk^51^. It is also possible that regulatory mutations contribute, perhaps modulated by folic acid.

Lastly, evidence of gene dosage sensitivity raises the possibility that environmental factors during critical developmental windows might amplify the effects of mutations.

## Methods

### Establishment of Spina Bifida Sequencing Consortium (SBSC) cohort

#### Inclusion criteria

Strict inclusion criteria were set for subject enrollment into the Spina Bifida Sequencing Consortium (SBSC). Participants must be affected by lumbosacral meningomyelocele (MM) with Arnold-Chiari malformation and hydrocephalus requiring surgical intervention such as ventriculoperitoneal shunt or endoscopic third ventriculostomy. We excluded subjects with closed neural tube defects, not requiring surgery at birth, with meningocele, or without hydrocephalus or not requiring surgery at birth. Enrollment required that both biological parents were available for sampling. In the case of fetal surgery to correct MM, which reduces the incidence of hydrocephalus, the requirement for inclusion of hydrocephalus at the time of enrollment was lifted. Subjects with known syndromes that would explain their conditions were excluded. Any families that did not meet the strict inclusion criteria were excluded.

#### Recruitment

The SBSC used several concurrent recruitment approaches: 1] Identify recruitable MM trios from local, national, and international hospitals. 2] Social media outreach through the Spina Bifida Association (SBA) Twitter, Facebook and Instagram accounts directly to families. 3] Recruit from spina bifida multidisciplinary clinics around the world with high MM caseloads. 4] Leverage historic Center for Disease Control-supported neural tube defect (NTD) cohorts. 5] Share sequencing data that have been already generated from trios with members of the SBSC and publicly with the NIH supported dbGaP. 6] Subjects and/or their families also contacted the SBSC independently through our postings and advertisements on social media. The countries where the participants were recruited include the United States, Mexico, Brazil, Italy, Georgia, Egypt, Canada, Venezuela, Pakistan, Guatemala, and Nigeria. All subjects or their guardians provided written informed consent approved by the UCSD Institutional Review Board S99075 protocol 140028, expiration date Aug 1, 2024. Recruitment processes were conducted in accordance with the approval by review boards of the University of California San Diego, operating under Federal-wide Assurance number, FWA00004495. The above rulings cover all aspects of our study, not just recruitment and consent.

#### Identifiers

The subject IDs used in this study cannot reveal the identity of the study subjects. These IDs are not know to anyone outside the research group.

#### Study questionnaire

All families prospectively recruited completed a standardized SBSC Questionnaire, which confirmed inclusion criteria, documented past medical history and current status by adopting prior questionnaire fields^52^, along with place and date of birth of affected and parents to control DNM rate for parental age at the time of conception.

#### Subject sample and data handling

DNA was extracted from blood or saliva by standard salt extraction protocols using Qiagen (Germantown, MD) or Autogen (Holliston, MA) proto\ prior CDC-funded recruitment efforts where recontact was not possible, and thus they were considered ‘lost to follow-up’ for sample re-collection. There were 37 families that had prior trio WES^15^ where data was incorporated into the cohort using the standardized bioinformatics pipeline.

### Sequence data generation

In total, 2541 samples from 851 MM trios (839 trios and 6 quartets) were eligible for whole exome sequencing with six target capture kits (Roche Exome V2, Agilent xGen Exome V1, Agilent SureSelect v4, Agilent SureSelect V4, IDT xGen Exome V2, and Twist human comprehensive exome), each trio sequenced by using the same capture kit. Samples with low concentration of DNA, gender discordant, or contamination yielded 45 samples to be failed quality control, resulting in 794 trios and 6 quartets. For control trios, we obtained 732 healthy trios from the SAFARI cohort^16^, 2202 WES data captured with NimbleGen EZ v2.

### Data preprocessing and quality control

Raw reads were aligned to reference genome (GRCh38) using bwa mem (0.7.17), preprocessed with PICARD (2.20.7) AddOrReplaceReadGroups and MarkDuplicates. Germline variants were collected with GATK (v4.11.0) HaplotypeCaller in the reference confidence model (-ERC GVCF) to be combined via CombineGVCF, generating a joint vcf for all MM and control trios, followed by GenotypeGVCF (v4.11.0) to be jointly genotyped. Multiallelic variants were splited (bcftools norm -m -any), Indels were realigned, and base quality recalibration was done with GATK (v.4.0.11) with known Indel and population frequency information (dbSNP (146) and Mills_and_1000G_gold_standard.indels.hg38.vcf.gz from GATK resource bundle).

Variant quality was recalibrated by GATK VQSR with HapMap (hapmap_3.3.hg38.vcf.gz), omni (1000G_omni2.5.hg38.vcf.gz), 1000G (1000G_phase1.snps.high_confidence.hg38.vcf.gz), (Mills_and_1000G_gold_standard.indels.hg38.vcf.gz), and axiom (Axiom_Exome_Plus.genotypes.all_populations.poly.hg38.vcf.gz).

To confirm the kinship integrity within trios, we used TRUFFLE^53^ (v1.38), removing 29 and 7 trios from MM and control cohort, respectively. IBD1 values were extracted from the vcf, by only using variants with allele frequencies higher than 0.05 (--maf 0.05) to only use confident calls. The IBD1 for all possible sample pairs were examined and if a parent-child IBD1 value is smaller than 0.75. Also, samples having more relations (IBD1 ≥ 0.75) with one outside of their family were removed, possibly owing to low data quality or contamination.

### De novo variant calling

De novo SNVs and Indels were collected following GATK genotype refinement steps. The genotypes of 777 MM and 725 control trios were gathered, and posterior probabilities of the variants were calculated with CalculateGenotypePosteriors (v4.2.6.1). We only selected de novo variants having genotype qualities (GQ) ≥ 20, Mapping quality (MQ) ≥ 30, and PASS in VQSR. Candidate DNMs that were found to have sequencing depth (DP) ≥ 12 in all three family members were kept to secure high-confident calls. To remove candidates that deviate from theoretical heterozygous states (AF=0.5), a two-sided binomial test was conducted to all positions by applying read depths and number of alternative alleles, removing sites with *P* value > 0.01. Genotypes of both parents were examined and kept if genotyped as reference homozygous (GT=0/0). To enhance the robustness of a high-confident set of DNMs, a series of post-filters were applied, by examining regional information and raw allele counts. First, clustered DNMs were filtered out when two or more DNMs of one proband were observed within 10 bp.

Second, DNMs that were found to exist in the same genotypes in parents either in MM or control cohorts were also removed. For de novo SNVs, each DNM position was examined by collecting raw reads with base quality (> 13) and mapping quality (> 0) criteria for each trio (bcftools mpileup v1.9). Then, to remove artifacts derived from miscellaneous alignment, a position was filtered out when a same alternative allele was observed in either of the parents. Two-sided binomial test was once more applied for SNVs with raw read counts, with a strict cutoff 0.05. All Indels were manually inspected with IGV (Integrative Genomics Viewer) by three experienced experts. The schematic overview of the DNM detection pipeline is shown in **Extended Data Fig. 16a**. VEP (v106) was used to annotate de novo variants with 1000 genomes (phase 3), COSMIC (92), ClinVar (202109), ESP (V2-SSA136), HGMD-PUBLIC (20204), dbSNP (154), GENCODE (Human Release 40), gnomAD (r2.1.1), PolyPhen (2.2.2) and SIFT (5.2.2). Mutational signatures were collected by Mutalisk^54^ using linear regression.

### DNM statistical confirmation

#### Consensus region for DNM rate calculation

To compare DNM rates and burden among sequencing data generated with different platforms (e.g., exome capture libraries), genomic regions that are commonly covered with sufficient coverage (that is, consensus region) were extracted. For each individual genome coverage was calculated using bedtools^55^ (v 2.30.0) genomecov. Then, the coverage bedgraphs were merged into each trio using bedtools unionbedg, and only the regions with read depth ≥ 12 in all three family members were kept. Each family bedgraph was merged with bedgraphs of other families within each batch, to confirm if at least 70% of families covered that region. Finally, the batch bedgraphs were merged into a final consensus region, reaching 36,553,428 bp.

#### DNM rates examined with Poisson distribution

DNMs located in the consensus region were collected, to confirm if the number of DNM per proband follows an expected Poisson distribution. Briefly, we assumed a cutoff of DNM count per proband from 1 to 20, generating DNM sets based on the cutoffs. Then, an expected Poisson distribution was generated per cutoff, with lambda as an average count of DNMs per proband (R dpoi). An observed distribution of DNMs per proband was calculated to be compared to the expected distribution, with Chi-Square goodness-of-fit test (R chisq.test). A cutoff of 7 DNMs per proband was set (**Extended Data Fig. 2c– 2d**), removing five and one trios from MM and controls, respectively. The six samples with DNM counts larger than 7 were found to have DNMs ranging from 8 to 484, possibly owing to low data quality.

### DNM burden analysis

DNM rates were calculated based on the number of DNMs observed in the consensus region, to properly compare the rates. We utilized a rate ratio test given that the number of DNMs per proband followed Poisson distribution. The DNMs were categorized into likely gene disrupting (LGD) including frameshift Indels, stop gained, and splice donor/acceptor mutations. Among the missense variants, if annotated as ‘probably damaging’ or ‘likely damaging’ in PolyPhen or ‘deleterious’ in SIFT, when CADD prediction score (>20) supports either at the same time, we annotated DNMs as damaging missense (D-Mis). If a DNM was annotated as D (deleterious) by a highly strict meta predictor MetaSVM^56^, we prioritized them as D-Mis-HC. We could replicate the highly consensus D-Mis mutations and their burden with two other independent missense variant annotation predictors, MetaLR^56^ and REVEL^57^ (**Supplementary Information** and **Extended Data Fig. 16b**). MetaSVM, MetalLR, and REVEL was annotated based on dbNSFP (4.1a).Missense mutations that were not annotated as D-Mis were categorized as ‘Tolerant missense’. Constrained genes were collected with pLI ≥ 0.9.

DNM rate was calculated within the consensus region (36,553,428 bp) in the mutation categories, with 95% confidence intervals calculated (one sample t test). Two-sided Wilcoxon rank-sum test was utilized to confirm the difference of the total DNM rates between MM (*n* = 772) and controls (*n* = 724). Rate ratio test (one-sided) was used to compare two Poisson distributed rates from each cohort. Theoretical DNM rates were estimated based on the total size of the coding region in hg38 (59,281,518 bp). The percentage of the subjects with DNM mediating risk could be calculated by the difference between the theoretical rates (DNM rate per exome) of the MM and control (95% CI calculated with Wilcoxon rank-sum test, two-sided). The percentage of DNM carrying the MM risk was estimated by dividing the difference of theoretical rate by the theoretical rate of MM trios.

### Whole genome analysis

We additionally recruited 101 trios and 1 quartet with MM and generated whole genome sequencing (WGS) data with Illumina HiSeq2500 and NovaSeq. The reads were preprocessed and aligned with the same pipeline of WES data. Same quality controls with WES data were applied, removing 4 trios that failed kinship integrity and contamination analyses. Small SNVs and Indels were collected with the same variant calling and filtering of WES analysis. We applied a binomial test to confirm if a DNM was derived from a heterozygous state as in WES, with applied stronger *P* value cutoff 0.05 compared to WES, as WGS data has much stable coverage with reduced capture bias and allelic imbalance. Additional populational allele frequency information (gnomAD genome v3.1.2) was added and SpliceAI (v1.3.1)^58^ was utilized (score > 0.5) to predict splice disrupting variants. We annotated noncoding variants with GREEN-VARAN^59^ (GREEN-DB schema 2.5) to annotate DNMs in noncoding regions. For copy number variant analysis (CNV), CNVpytor (1.3.1)^60^ was utilized with a window size of 100 kbp, and manually curated by confirming the read depths of all father and mother. To comprehensively detect structural variants (SVs), we utilized Manta^61^ (1.6.0), Delly^62^ (v0.8.1), and smoove^63^ (0.2.6) and merged the SV calls from the three callers with Survivor (1.0.7, parameter: dist=50000, callers=0, type=0, strands=0, estimate=0, size=50000) to confirm if the detected SVs only present in probands.

### Multiplexed Error Robust In-Situ Hybridization (MERFISH)

MERFISH was performed as described previously at the UCSD Epigenomics Core^64^. Briefly, mouse embryos at day 10.5 were fixed in paraformaldehyde and cryopreserved in 30% sucrose, embedded in Optimal cutting temperature (OCT) and sectioned into 12 μm sections in cryostat. These tissue sections then underwent fluorescent in situ hybridization with a panel of oligonucleotide probes specific for 36 of the genes identified from the current human DNM cohort along with 107 marker genes, chosen upon previously published single cell literatures to classify cell types into 7 cell types (neuron, neural progenitor, neural crest, pre-epithelial to mesenchymal transition neural crest progenitor (Pre-EMT-NCP), dorsal root ganglia, blood and mesoderm^65–67^. During downstream analysis marker genes were filtered to only include 66 the most efficient markers specific for each cell type (see **Supplementary Data** for a full list of utilized marker genes).

Probes were designed and manufactured with standard pipeline by Vizgen (https://vizgen.com/gene-panel/). Tissue sections were prepared and processed using Vizgen sample preparation protocol (Document Number 91600002 Rev A), then samples placed in the Merscope for imaging and decoding (Document Number 91600001, Rev G). Raw data was analyzed using Scanpy (1.9.1)^68^. Cells were preprocessed and filtered using the following criteria: cells with volume less than 100 μm^3^, cells with volume larger than 3x median volume of all cells, fewer than 4 genes detected, fewer than 10 transcripts detected, total RNA counts lower than the 2% quantile or higher than 98% quantile. Preprocessed cells were clustered based on gene expression using the Leiden algorithm (leidenalg 0.10.2), annotated using scType^73^ (v1.0) based upon marker genes as the reference set (see **Supplementary Information**). Clusters were annotated to the cell type with the highest overall score from scType, accounting for expression of all marker genes per cell type in a focal cluster, compared to the expression in other clusters.

Clusters with prediction score less than 0 or low confidence scores (score less than 10% of the number of cells in each cluster) were classified as indeterminate as they could not be classified in any of the 7 cell types. Expression of each of the 36 candidate genes was then analyzed in each cell from raw data, and percentage of total average expression per cell type calculated in **Extended Data Fig. 5**.

### Network analysis (co-localization and degree)

Relationship between damaging DNMs in MM vs. control cohorts and the human genome network were calculated using STRING (v11.5) for gene interactions. We calculated the connectedness from the STRING database of damaging DNMs from MM (*n* = 187) and control (*n* = 135) among 19,699 annotated genes, randomly resampled with a size of 80% of the gene set from control (*n* = 108), of 100,000 iterations. Number of edges per iteration were compared with a Wilcoxon rank-sum test (two-sided). Edges per nodes were compared between the cohorts (Wilcoxon rank-sum test, one-sided).

### Pathway enrichment analysis

To estimate if the MM or control DNM gene sets were functionally enriched in known biological pathways, gene ontology (GO) biological process, reactome, and KEGG pathways, we conducted a gene enrichment analysis with ClueGO (v2.5.10) in Cytoscape (3.10.1), with parameters set as Evidence All_without_IEA (Inferred from Electronic annotation), GO tree interval ranging from 3 to 8, and 5 minimum number of genes, 4% of genes for GO term selection, and 0.4 kappa score. We used adjusted *P* value with Bonferroni step-down with significance criterion 0.05. Detailed information of the GO enrichment test can be found in **Extended Data Table 3**. No significant term was observed with control gene sets.

### Sanger confirmation

Out of 198 damaging DNMs, 86% (171 out of 198) where trio DNA samples were available for the Sanger sequencing confirmation, with sufficient DNA. For a subset of detected damaging DNMs, primer sets were generated via Primer3 and ordered from IDT. The primer lengths were configured to span a range of 18 bp to 23 bp, targeting an ideal size of 20 bp. The GC content was set to a minimum of 30% and a maximum of 70%, and the annealing temperature was established to range between 57°C and 62°C, with an optimal temperature of 59°C. PCR was performed using the QIAGEN Taq DNA Polymerase kit and primer concentrations of 500 nM. Four reactions were performed per DNM, amplifying DNA from the father, mother, affected child, and an unrelated healthy individual. PCR products were confirmed by agarose gel electrophoresis, purified by incubation with Exonuclease I/Shrimp Alkaline Phosphatase (1:2), diluted in ddH2O, and shipped to Genewiz/Azenta for Sanger sequencing. Trace files were analyzed in SnapGene Viewer and compared against the UCSC Genome Browser to confirm that the affected child was heterozygous for the detected mutation and the father, mother, and control were homozygous for the reference base at the same locus.

### Network propagation and submodule identification

A propagated network was constructed with NetColoc (0.1.7)^69^. PCNet^70^ was utilized for the background human gene interaction network, which contains 18,820 nodes and 2,693,109 edges. Among 187 MM genes, two genes (SLCO1B3-SLC01B7 and H3-4) couldn’t be included as seeds because they couldn’t be found in the PCNet. As implemented in NetColoc, w prime and w double prime value was calculated for each gene to generate z scores for proximity. We used the z score threshold 3 as default. The entire list of the genes in the propagated network can be found in **Supplementary Data**. Known mouse NTD genes (*n* = 374) were based on previous literatures, where 205 genes were organized back in 2010^30^ and the others were added from then (see **Supplementary Data** for the gene list). To examine the propagated network, we conducted a hypergeometric test, utilizing 18,820 as the total number of genes as in PCNet (*M* = 18,820), number of known NTD genes (*n* = 374), the number of nodes in the propagated network (*N* = 439), and the number of overlapping genes with propagated network and MM genes (*k* = 17).

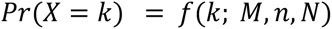

Then, to identify small functional modules from the expanded network, the propagated network was clustered with a Leiden algorithm with 0.5 resolution, 0.01 beta with 45 iterations. To use co-expression value as attributes, we again transferred the propagated network to STRING database (v12) with Homo Sapiens full STRING network, popping out 114 nodes without edges to the network, meaning discrepant sources of protein network. In total, 15 small clusters were generated with more than five nodes. Five could be significantly enriched by functional biological terms (FDR < 10^-5^) by the GO biological process, KEGG pathway without disease related terms, and Reactome, which had more than 30% of the nodes derived from damaging DNMs (**Supplementary Data)**. Two clusters annotated as ‘GTPase involved actin cytoskeleton’ and ‘Microtubule-based process’ were enriched with functional terms with most significant *P* values (FDR < 10^-8^), with 8 and 4 terms respectively. The pLI of the genes in the five functional modules can be found in **Supplementary Data**.

### Cloning

Patient DNMs were mutagenized into wildtype (WT) cDNA using Gibson Assembly cloning. These mutations include p.R231G (R230G in mouse) in *Vwa8b* (mouse), p.E521Q and p.E623Q in *PLCE1* (human), p.P168L in *TNK2*, p.H1149P *TIAM1,* p.R620H in *BRSK2* (human) and p.R349C in *CLIP2* (rat). For each of the plasmids, overlapping (40 bp) forward and reverse primers with 20 bp 5’ overhangs were designed complementary to the mutation site, with a single nucleobase difference corresponding to the mutation of interest (**Supplementary Data**). 15 ng plasmid DNA was amplified using the NEB Phusion High-Fidelity 2X Master Mix [NEB #M0531] for 25 cycles, after which the reaction was subjected to 1 hour at 50°C with NEBuilder HiFi DNA Assembly 2X Master Mix [NEB #E2621], and the reaction mixture was treated with *DpnI* [NEB #R0176] digestion for 37°C for 30 minutes. 2 µL of the reaction mixture was then transformed into NEB DH5-alpha Competent *E. coli* [NEB #C2987], with successful mutation verified using Primordium whole plasmid sequencing.

### Protein analysis

#### TNK2

##### Cell transfection, lysis, and Western blotting

Human embryonic kidney (HEK)293T cells (from American Type Culture Collection) were maintained in DMEM (Corning) supplemented with 10% fetal bovine serum (Sigma) and 1X penicillin and streptomycin. The cells were transfected according to the protocol supplied with TransIT reagent (Mirus) using 7.5 μg of DNA. After 48 hours, the cells were harvested, washed with phosphate-buffered saline, and lysed in a buffer containing 25 mm Tris–HCl (pH 7.5), 100 mm NaCl, 1 mm EDTA, 1% Nonidet P-40, 5 μg/ml leupeptin, 5 μg/ml aprotinin, 1mM PMSF, and 200 μM Na3VO4. Cell lysis proceeded for 30 minutes at 4°C with rocking. The cell suspensions were centrifuged at 15,000 rpm for 10 minutes, and protein concentrations of the soluble lysates were measured using a colorimetric Bradford assay. The lysates (100 µg) were analyzed by SDS-PAGE and Western blotting using the following antibodies: phospho-Ack1/Tnk2 (Millipore Sigma 09-142), Flag (Sigma-Aldrich A8592), and gamma-tubulin (Sigma-Aldrich T6557).

##### Immunoprecipitation (IP) Kinase assay

IP-kinase assays were performed essentially as described with the following modifications: HEK293T lysates (1 mg) were incubated with 40 μL of anti-Flag M2 affinity resin (Sigma) on a rotator at 4 °C overnight, then washed three times with Tris-buffered saline (TBS). The IPs were divided into three portions. One portion of each sample was mixed with a gel-loading buffer, boiled for 3 minutes, and analyzed by 7.5% SDS/PAGE with anti-Flag Western blotting. The remaining two samples were used for duplicate activity measurements using the phosphocellulose paper-binding assay. The reactions contained 20 mm Tris–HCl (pH 7.4), 10 mm MgCl2, 0.25 mm ATP, 1 mM WASP peptide (KVIYDFIEKKG) and 20–50 cpm/pmol of [γ-32P] ATP. Reactions were carried out at 30 °C for 15 min. Incorporation of 32P into peptide was measured by scintillation counting.

#### TIAM1

##### DNA Reagents

The following constructs were previously described: pCMV-EGFP^71^, pCMV-Flag-Tiam1 (WT-Tiam1)^72^, pRaichu-RaichuEV-Rac1 (2248X)^73^, pcDNA3.1 vector^74^. mRuby3 was obtained from Addgene (127808).

##### Cell culture and transfections

Cos-7 cells were grown in Dulbecco’s modified Eagle’s medium (DMEM) (Corning 10-013-CV) supplemented with 100 U/ml penicillin/streptomycin (Invitrogen 15140122) and 10% heat inactivated fetal bovine serum (Atlanta Biologicals S11150H) on cell culture-treated plastic (VWR 10062-880) maintained at 5% CO2, 37°C. Cells were passaged with trypsin (Invitrogen 25200072) once per week. For experiments, 24 hrs prior to transfection cells were plated on either nitric acid washed glass cover slips (Bellco Glass 1943-10012A) or glass bottom plates (Cellvis P241.5HN) coated with 20 ug/ml Poly-D-Lysine (Corning 354210). Cells were transfected using Jetprime reagent (Polyplus 101000046) according to the manufacturer recommendations. Twenty-four hours after transfections cells were changed to serum-free DMEM. For live cell experiments, phenol-red free DMEM (Corning 17-205-CV) was used. Experiments were conducted 48 hours after transfection.

##### Immunocytochemistry, microscopy, and analysis

Cells were fixed with 4% Paraformaldehyde (Fisher Scientific AC169650010), 4% Sucrose (Sigma S0389) in phosphate buffered saline (PBS). To confirm construct expression, following fixation all cells were immunostained overnight at 4°C with anti-Flag (Cell Signaling 14793S) primary antibody diluted in 0.3% Triton X-100, 5% goat serum in PBS. After, cells were incubated with Cy5 anti-Rabbit secondary antibody (Jackson ImmunoResearch 111-175-003) for 2 hrs at room temperature. For filamentous actin experiments, cells were also incubated with Texas Red-Phalloidin (Invitrogen T7471) during this time. Post-fixation and staining, cells grown in glass coverslips were mounted using the aqueous mounting solution FluorSave (EMD Millipore 345789).

##### Imaging

Filamentous actin was labeled with Texas Red phalloidin, then imaged using a Zeiss AxioObserver.Z1 microscope with a 20x objective. Rac1 activation was measure in cells transfected with the RaichuEV-Rac1 probe, imaged on a Zeiss LSM 880 confocal microscope at 20x. Live imaging experiments were conducted at 37°C/5% CO2. Single plane images were acquired with Förster resonance energy transfer (FRET) (excitation 458 nm, emission 522–569 nm), CFP (Donor) (excitation 458 nm, emission 463–507 nm), and YFP (Acceptor) (excitation 514 nm, emission 522–569 nm), presented as normalized FRET/Donor values using ImageJ (NIH). Only cells expressing constructs were selected for analysis. GFP or mRuby3 fill visualized cell morphology via ROI cell trace, measured for mean intensity, corrected for background, and normalized to respective controls. Experiments were conducted with experimenters blinded to conditions. Representative images were masked using ImageJ (NIH).

#### VWA8

Plasmid transfection in HEK293T cells was performed using the ThermoFisher Lipofectamine 2000 transfection reagent. HEK293T cells seeded on a 6 well plate and transfected with 4ug of plasmid. Cells were lysed using Modified RIPA buffer 72 days post-transfection, and the lysate was spun down at 10,000g in a microcentrifuge and supernatant was harvested. Empty vector, WT, and R230G protein was then quantified using the Pierce BCA Assay kit (Thermo Scientific #23225), and 10 µg protein lysate was diluted in 4X SDS-PAGE Loading Buffer supplemented with 10% β-mercaptoethanol (Sigma #M3148) and loaded into each well of an Any kDa Bio-Rad Mini-PROTEAN TGX [4569031] gel, and run for 30 minutes at 150 V. Proteins were then transferred onto a nitrocellulose membrane [Sigma #IPVH00010] and blocked with 5% BSA (Sigma #A9418) in TBST (blocking buffer) for 1 hour at room temperature. The membrane was then blotted with 1:1000 Anti-HA (#sc-805) and 1:5000 Anti-alpha-tubulin (Sigma #T6074) primary antibody in blocking buffer overnight at 4℃ under constant rocking. The following day the membrane was washed 4 times in TBST for 10 minutes, followed by1 hour incubation in a 1:10000 anti-mouse (#715-035-150) and anti-rabbit (#711-036-152) HRP-conjugated secondary antibody in blocking buffer at room temperature. The membrane was then washed again for 4 times in TBST for 10 minutes, after which the membrane was treated with Pierce ECLWestern solution for HRP detection (Thermo Scientific #32106) for 5 minutes at room temperature. The membrane was then transferred into a clear plastic cover and imaged using a chemiluminescence detector (Biotechnique #92-14860). Quantification of protein band intensity was accomplished using the ImageJ band densitometry plugin, and final plots were created using Prism.

#### CLIP2

##### Western blot

HEK293T cells were transfected with pEGFP-CLIP2 WT, R349C (TGC or TGT) either with constitutively active AKT pE17K (Addgene #73050) or empty vector using Lipofectamine as previously described, lysed into sample buffer, and analyzed by western using anti-GFP antibody (1:200 Thermo Scientific #GF28R).

##### Microtubule analysis

HuH7 cells cultured in DMEM with 10% fetal calf serum (Sigma) and 1% antibiotic-antifungal mixture (Gibco) were transfected with pEGFP-CLIP2 WT, R349C or S352A (Turbofect, ThermoFisher Scientific #R0531). Time-lapse sequences (Δt = 2.4 s, 80 frames) of the CLIP2 fluorescence signal were captured using a Leica DMLB microscope through a 100x 1.3 NA objective and a Scion CFW1312M camera. The parameters of microtubule dynamics were computed as previously described^75^. Statistical comparisons were performed using one-factor ANOVA.

#### PLCE1

##### Cell transfection and lysis

Plasmid expressing PLCE1 including the WT, p.E521Q, and p.E623Q mutations were transfected into HEK293 cells and were maintained in DMEM supplemented with 10% FBS and 1% penicillin/streptomycin. Plasmids were transfected into HEK293 cells using Lipofectamine (Invitrogen), grown for 48 hr and lysed in lysis buffer (1% Triton X-100, 50 mM Tris pH 7.4, 10 mM MgCl2, 500 NaCl) with added protease inhibitors (Roche). The lysates were spun down at 20000 g/4°C/15 min and the supernatants were used for overnight incubation with 20 μl of GST beads containing GST recombinant proteins on the rotator. After the incubation samples were washed with 800 μl lysis buffer, the GST beads were collected at 500 g/4°C/2 min, the supernatants were aspirated out and this washing was repeated 3 times. Proteins were eluted with 20 μl of 2x Laemmli buffer.

##### Pulldown assay

Eluted proteins were diluted in 4X SDS-PAGE Loading Buffer supplemented with 10% β-mercaptoethanol (Sigma #M3148) and loaded into each well of an Any kDa Bio-Rad Mini-PROTEAN TGX gel, and run for 30 minutes at 150 V, samples transferred to PVDF membrane (Sigma #IPVH00010), blocked with 5% BSA (Sigma #A9418) in TBST for 1h at RT, membrane blotted with 1:1000 anti-RAC1, 1:1000 anti-CDC42, 1:1000 anti-RHOA, and 1:1000 anti-Myc, washed in TBST for 10min then blotted with 1:10000 anti-mouse (#715-035-150) and anti-rabbit (#711-036-152) HRP-conjugated antibody, washed in TBST for 10m, then developed in Pierce ECL Western solution (Thermo Scientific #32106) for 5min, and imaged using a chemiluminescence detector [Biotechnique #92-14860], and band intensity quantified using ImageJ band densitometry, and plotted using Prism.

The GST-PAK1 beads (#PAK02-A) and GST-rhotekin RBD beads (#RT02-A) were purchased from Cytoskeleton. Anti-Myc (#2278; Cell Signaling Tech.), anti-RhoA (#2117; Cell Signaling Tech.), anti-Rac1 (#610650; BD Transduction Labs), anti-Cdc42 (#610928; BD Transduction Labs) were purchased from the indicated commercial sources.

#### BRSK2 Western Blot

HEK293T cells were transfected with the pEXP-CMV-BRSK2-Flag plasmid containing wildtype, K48A kinase-dead mutant, and R620H patient mutation BRSK2 sequence using PEI (Sigma Millipore #919012) and OptiMEM (Gibco #31985-070) according to the manufacturer’s guidelines. Protein concentration of lysates was measured using the Pierce BCA Protein Assay Kit (Thermo Scientific #23225), then 20 µg of protein was loaded into each well of a 4-12% Bis-Tris Gel (Invitrogen #NP0336), transferred to nitrocellulose membranes (Thermo, #88018), blocked then blotted with one of the following primary antibodies: 1:1000 Flag rabbit (Cell Signaling Technology #14793S), 1:1000 Beta-actin rabbit (Sigma #A5316), 1:1000 anti-Phospho-AMPK Substrate Motif rabbit (Cell Signaling Technology #5759) and 1:1000 mTOR rabbit (Cell Signaling Technology #2983), 1:1000 P-mTOR Ser 2448 rabbit (Cell Signaling Technology #5536), 1:1000 p70 S6 Kinase rabbit (Cell Signaling Technology #34475), 1:1000 Phospho-p70 S6 Kinase Thr 389 rabbit (Cell Signaling Technology #9234), 1:1000 4E-BP1 rabbit (Cell Signaling Technology #9644) and 1:1000 Phospho-4E-BP1 Thr 37/46 rabbit (Cell Signaling Technology #2855) at 4°C overnight, rinsed in TBST for 10 minutes, incubated with LI-COR IRDye 680 Donkey anti-Mouse or 800 Donkey anti-Rabbit (LI-COR #926-68072, #926-32213) 1:10,000 rinsed in TBST and imaged on an Odyssey CLx using Image Studio software for analysis of band intensity.

#### *Xenopus* modeling

*Xenopus* embryo manipulations were conducted according to established protocols^76^. Female adult *Xenopus laevis* were induced to ovulate by the injection of human chorionic gonadotropin, and *in vitro* fertilization was performed by homogenizing a small piece of a testis. Embryos were dejellied in 1/3 X Marc’s Modified Ringer’s (MMR) with 2.5% (w/v) cysteine (pH7.9) at the 2-cell stage and were microinjected with either gene-specific MO or mock into two dorsal blastomeres at the 4-cell stage in 2% Ficoll (w/v) in 1/3 X MMR. Injected embryos were incubated at 18℃ until stage 19 and fixed with 1X MEMFA (0.1 M MOPS, 2mM EGTA, 1 mM MgSO4, 3.7% formaldehyde, pH 7.4). The gene sequence for *Xenopus* Whamm was obtained from Xenbase (http://www.xenbase.org). A Morpholino antisense oligonucleotide (MO) against Whamm was designed to block mRNA splicing (Gene Tools) 5’ – AAAAGTAGGAAGAAGCCCCCACCCT -3’. The Whamm open reading frame was amplified from *Xenopus* cDNA and cloned into the pCS10R MCC vector containing GFP tag. Capped mRNA was synthesized using the mMESSAGE mMACHINE SP6 transcription kit (Invitrogen Ambion, cat# AM1340), and injected to assess rescue. To measure the distance of the neural folds, *in situ* hybridization against Pax3 was performed as previously described^76^. Images were captured using a ZEISS Axio Zoom V16 stereomicroscope and associated Zen software, and neural folds visualized by pax3 were analyzed in Fiji. To verify the efficiency of MO, MO was injected into all cells at the 4-cell stage, and total RNAs were extracted from four embryos using the TRIZOL reagent (Invitrogen, cat # 15596026). cDNAs were synthesized using M-MLV reverse Transcriptase (Invitrogen, cat#28025013) and random primers (NEB, cat#S1330S). cDNAs were amplified by Taq polymerase (NEB, cat# M02735) using the following primers: 5’-CGGTGCAGGACTTGGATTAT-3’, 5’-CCCATTTAGCTACCTCCTTCTG-3’.

## Data Availability

Exome and genome sequencing data are available in publicly accessible databases for the 1,576 subjects enrolled after inclusion of dbGaP language into the consenting process/ became standard. These data are at dbGaP (accession #phs002591.v1.p1, phs000744.v5.p2 (Yale Mendelian Sequencing Center), Kids First: Whole Genome Sequencing in Structural Defects of the Neural Tube Accession (phs002591.v1.p1).

Sequencing data on the remaining 743 subjects is available upon request.

## Code Availability

Computational codes used in this study are available in GitHub (https://github.com/Gleeson-Lab/Publications/tree/main/MM_DNM).

## Acknowledgement

We especially thank the families and the individuals with meningomyelocele that participated in this study, Kiely James, Renee George, Brett Copelande, Valentina Stanley, Celine Shen, and Jennifer Venneri from the Spina Bifida Sequencing Consortium, the Spina Bifida Association, the UCSD Laboratory for Pediatric Brain Disease for clinical and technical support, Brin Rosenthal and Katie Fisch from the UCSD Altman Clinical and Translational Research Institute for statistical modeling, Broad Institute, Yale Genetic Center, Regeneron Genetics Center, UCSD Institute for Genomic Medicine, UC Irvine Sequencing Center and Rady Children Institute for Genomics Medicine for sequencing support, Barbara Craddock from Stony Brook University for functional analysis of TNK2, and the Spina Bifida Association for recruitment efforts.

## Spina Bifida Sequencing Consortium members

Dr. Allison Elizabeth Ashley Koch, PhD (Duke University), Dr. Kit Sing Au, PhD and Dr. Hope Northrup, MD (University of Texas, Houston), Dr. Gyang Markus Bot, MD (Jos University Teaching Hospital, Jos, Nigeria), Valeria Capra, MD (IRCCS Istituto Giannina Gaslini, Italy), Dr. Richard H. Finnell, PhD, DABMGG (Baylor College of Medicine), Dr. Zoha Kibar, PhD (Université de Montréal, Canada), Dr. Philip J. Lupo, PhD, MPH (Baylor College of Medicine), Dr. Helio R. Machado, MD (University of Sao Paulo, Brazil), Dr. Tony Magana, MD (Mekelle University, Mekelle, Ethiopia), Dr. Rony Marwan, MD (University of Colorado), Dr. Gia Melikishvili, MD (MediClub Georgia, T’bilisi, Georgia), Dr. Osvaldo M. Mutchinick, MD, PhD (Salvador Zubirán National Institute of Health Sciences and Nutrition, Mexico City, Mexico), Dr. Roger E. Stevenson, MD (Greenwood Genetic Center), Dr. Anna Yurrita, MD (Francisco Marroquín University, Guatemala City, Guatemala), Dr. Maha S. Zaki, MD, PhD (National Research Center, Cairo, Egypt), Dr. Sara Mumtaz, PhD (National University of Medical Sciences, Rawalpindi, Pakistan), Dr. José Ramón Medina-Bereciartu, MD (Venezuela Association of Spina Bifida, Caracas, Venezuela), Dr. Friedhelm Hildebrandt (Boston Children’s Hospital, Harvard Medical School), Dr. Caroline M. Kolvenbach, MD (Boston Children’s Hospital, Harvard Medical School), Shirlee Shril, MS (Boston Children’s Hospital, Harvard Medical School), Dr. Mahmoud M. Noureldeen, MD and Dr. Aida MS. Salem, MD (Beni-Suef University, Beni-Suef, Egypt), Dr. Joseph Gleeson, MD (UC San Diego).

## Author Contributions

Y-JH, CW, NM FJ, KIV, CB, SL, NJ, AP recruited subjects and performed genetic analysis. IJ performed MERFISH analysis, CJL and JBW generated Xenopus data. AN, J-EL, IT, FAB, KFT, SY, HJ, BB WTM, CH, SAM, HYG, BB, CP performed functional analysis, ZK GMB, HN, KSU, MS, AA-K, RHF, JL, HM, CA, HRM, RES, AY, SM, OMM, JRM-B, FH, GM, RM, VC, MMN, AMSS, MYI, MSZ recruited families. AA, ARS, SWK, performed sequencing. Y-JH, SK and JGG performed analysis, wrote drafts, and incorporated feedback from coauthors.

## Additional Information

Supplementary Information is available, containing Supplementary Table and Supplementary Notes.

## Extended Data Figure

**Extended Data Figure 1.**
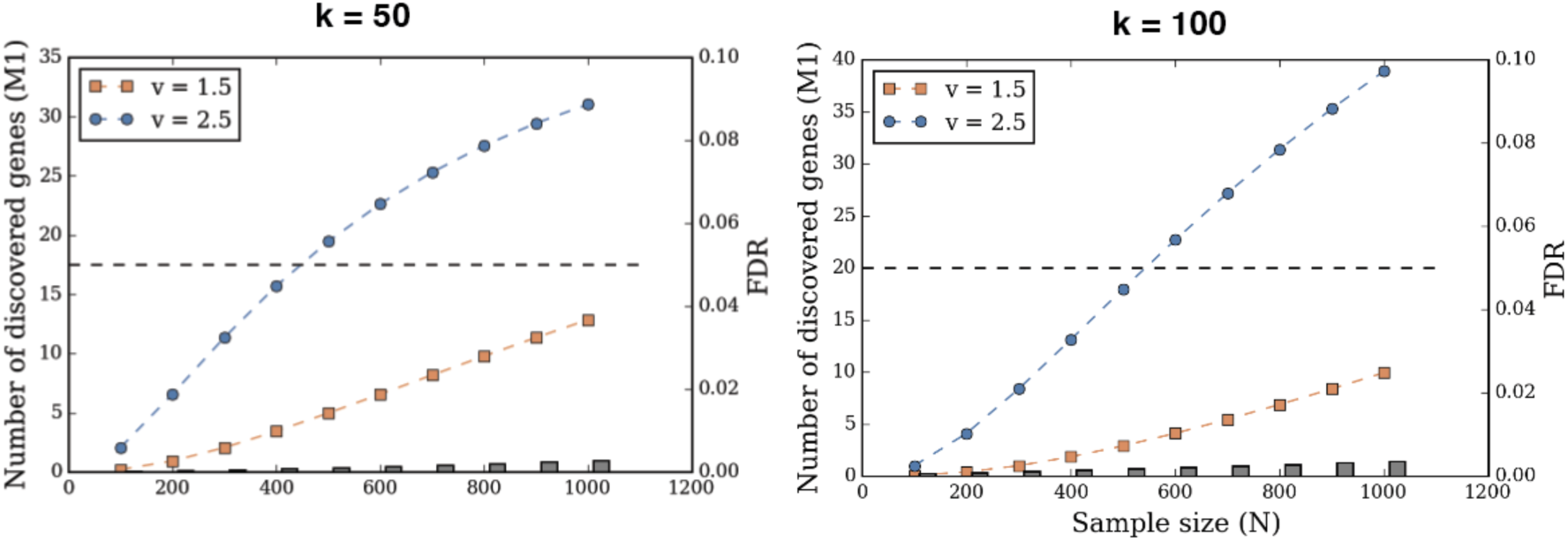
Power calculation of estimating a cohort size for DNM detection. Power calculation showing potential number of discovered genes compared with cohort size (350 trios), for two different v values (enrichment ratio of loss of function variants in case versus control) and two different k values (assumed number of risk genes). For instance, if there are 50 genes to discover (k=50), a cohort of 400 trios will identify 16 genes if LOF variants are 2.5x more common in affected (v=2.5). All calculations manage a conservative false discovery rate (FDR). Gray dash: FDR.

**Extended Data Figure 2.**
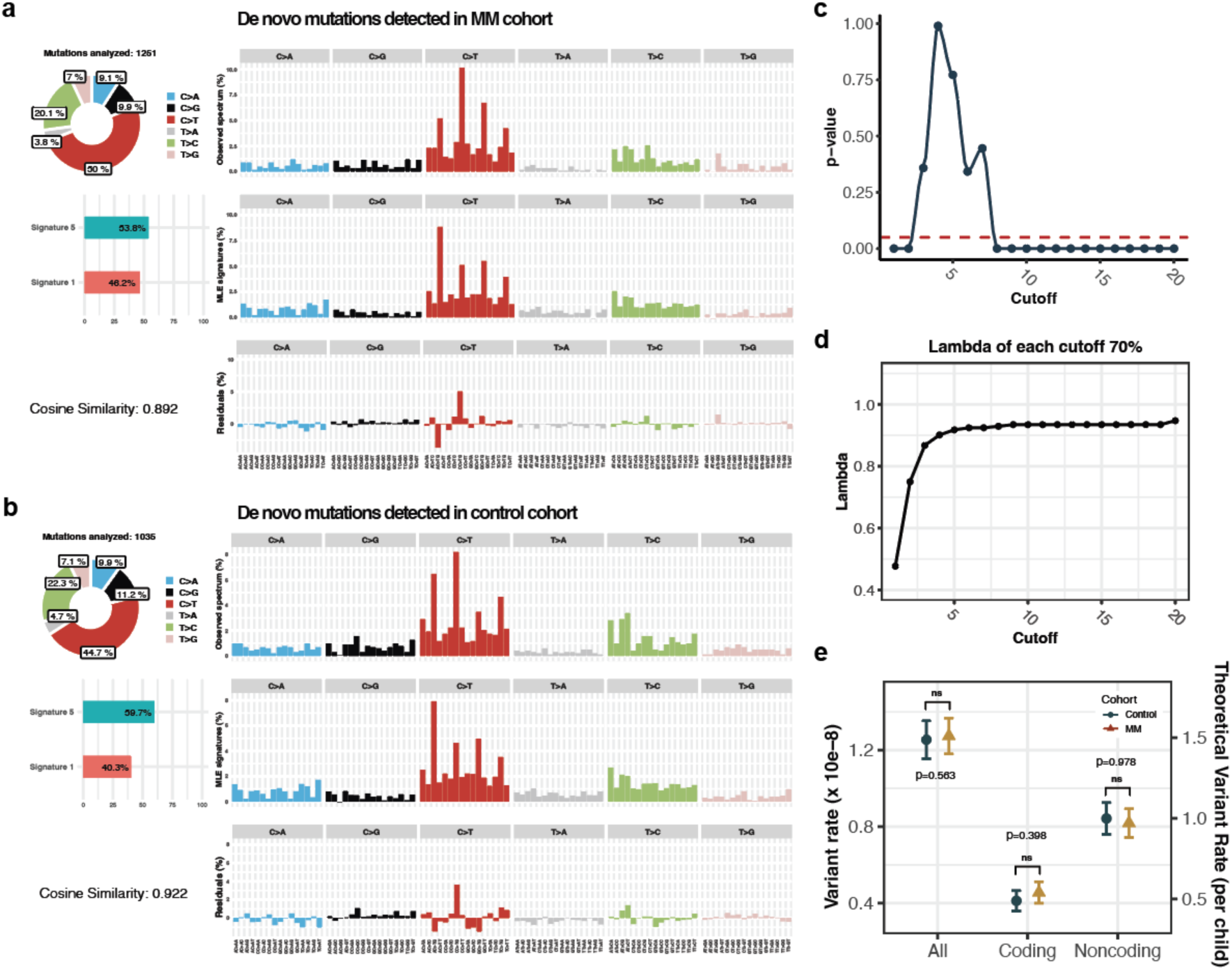
Quantitative and qualitative features of de novo variants. **a-b**, Mutational signatures of DNMs showing known de novo signatures 1 and 5, in both (**a**) MM and (**b**) control cohorts. 1,251 and 1,035 SNVs were utilized for the signature analysis. **c-d**, DNM counts per proband following Poisson distribution, examined based on cutoffs (DNM count per proband) ranging from 1 to 20. (**c**) Chi-Square goodness-of-fit (two sided) was performed with an expected Poisson distribution with (**d**) lambda as an average DNM count per proband, for all cutoffs (see **Methods**). **e**, DNM rates for all, coding, and noncoding regions. Observed and theoretical DNM rates are marked in the left and right y-axis, respectively. Theoretical rate calculated by normalization of the variant rate with the total size of hg38 coding region (59,281,518 bp). *P* values were calculated by a two-sided Wilcoxon rank-sum test. ns: not significant.

**Extended Data Figure 3.**
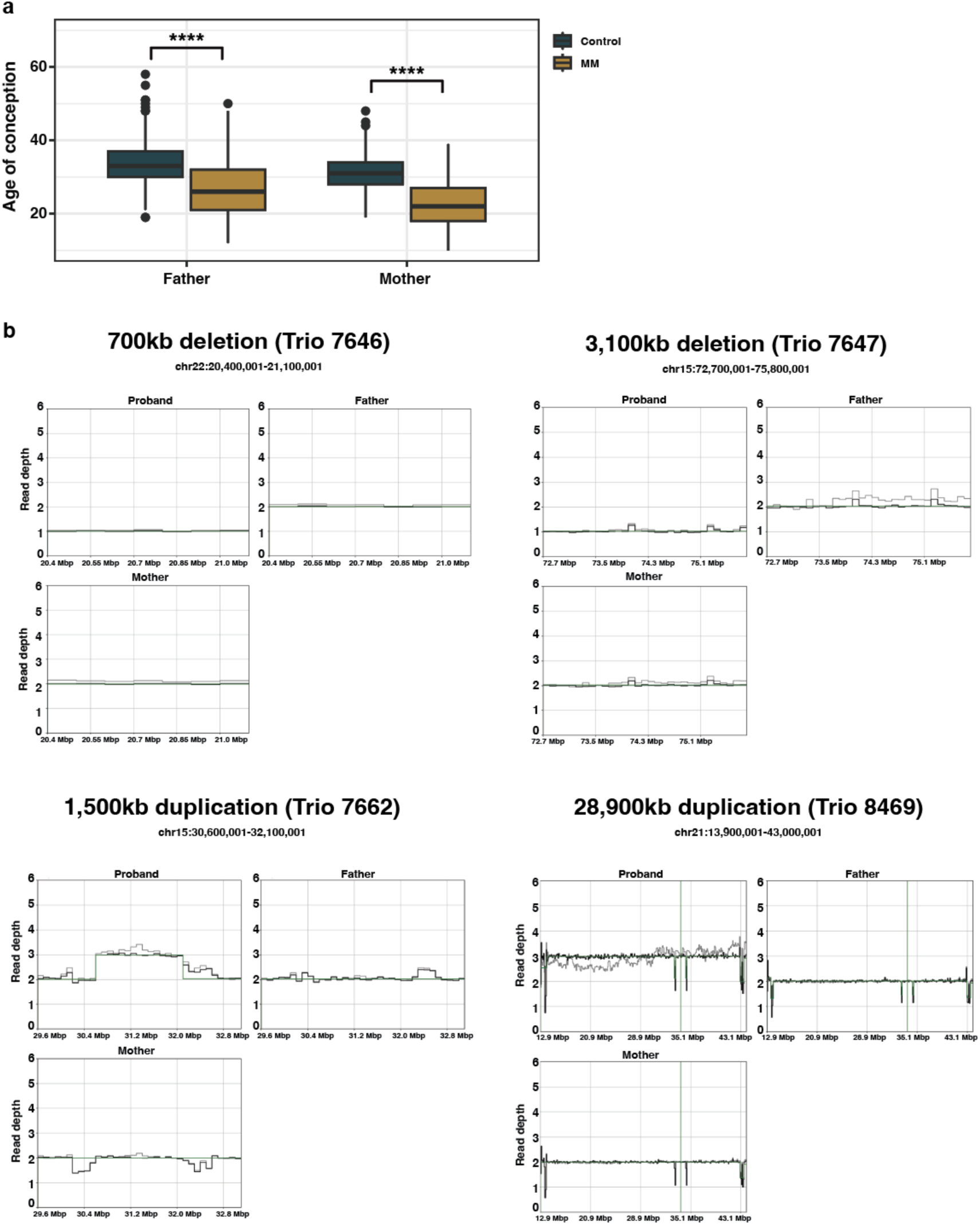
Quantitative and qualitative features of de novo variants. **a**, Age of conception of father and mother in MM (*n* = 79) and control (*n* = 683) trios. The ages of conception were compared with Wilcoxon rank-sum test (two sided). The age of conceptions were significantly higher in controls than MM, for both of the parents. *P* values: **** < 0.0001. **b**, De novo large CNVs (> 100kb) detected from WGS. Read depths of proband, father, and mother shown. Gray lines: read depths. Green lines: copy number.

**Extended Data Figure 4.**
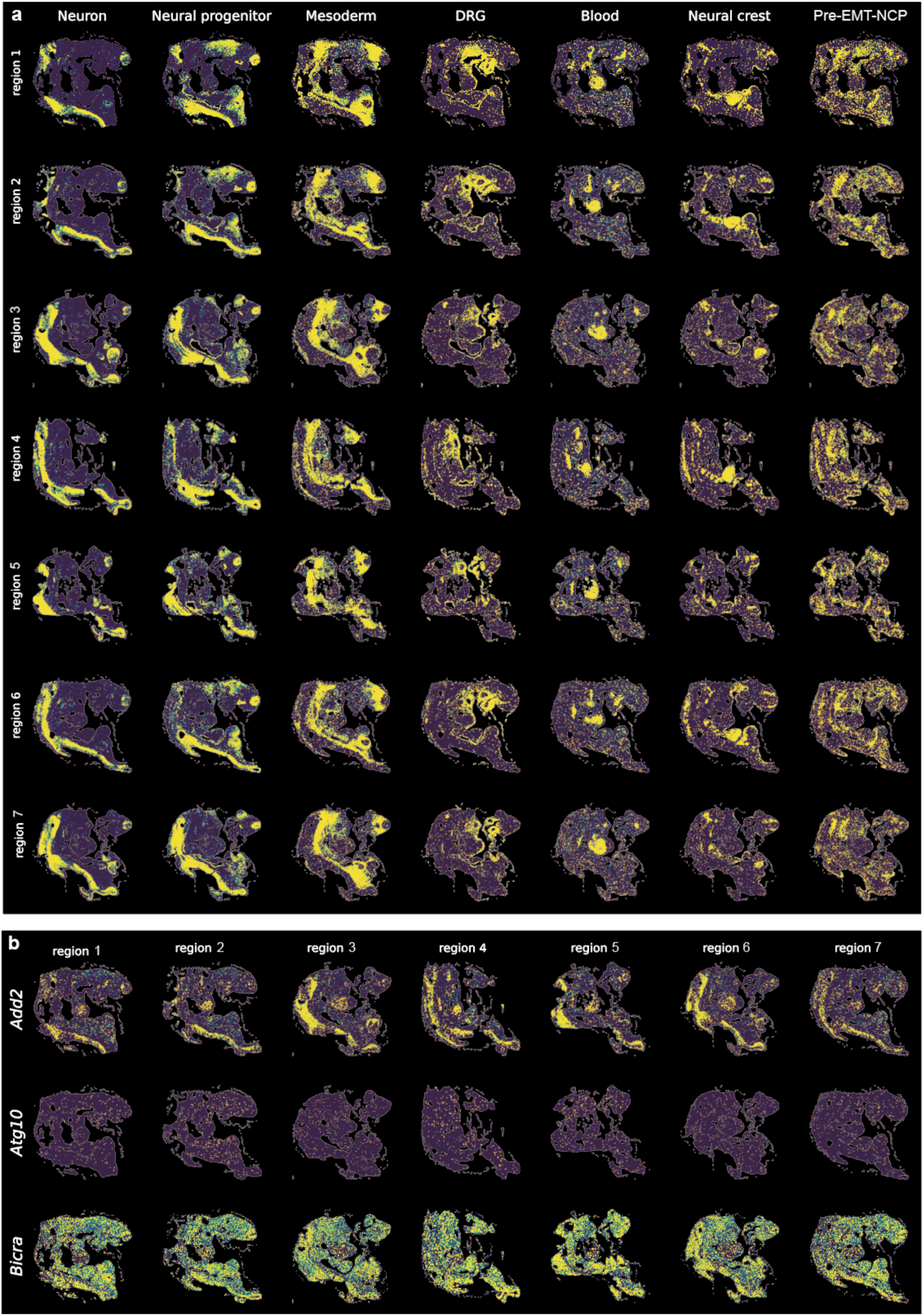
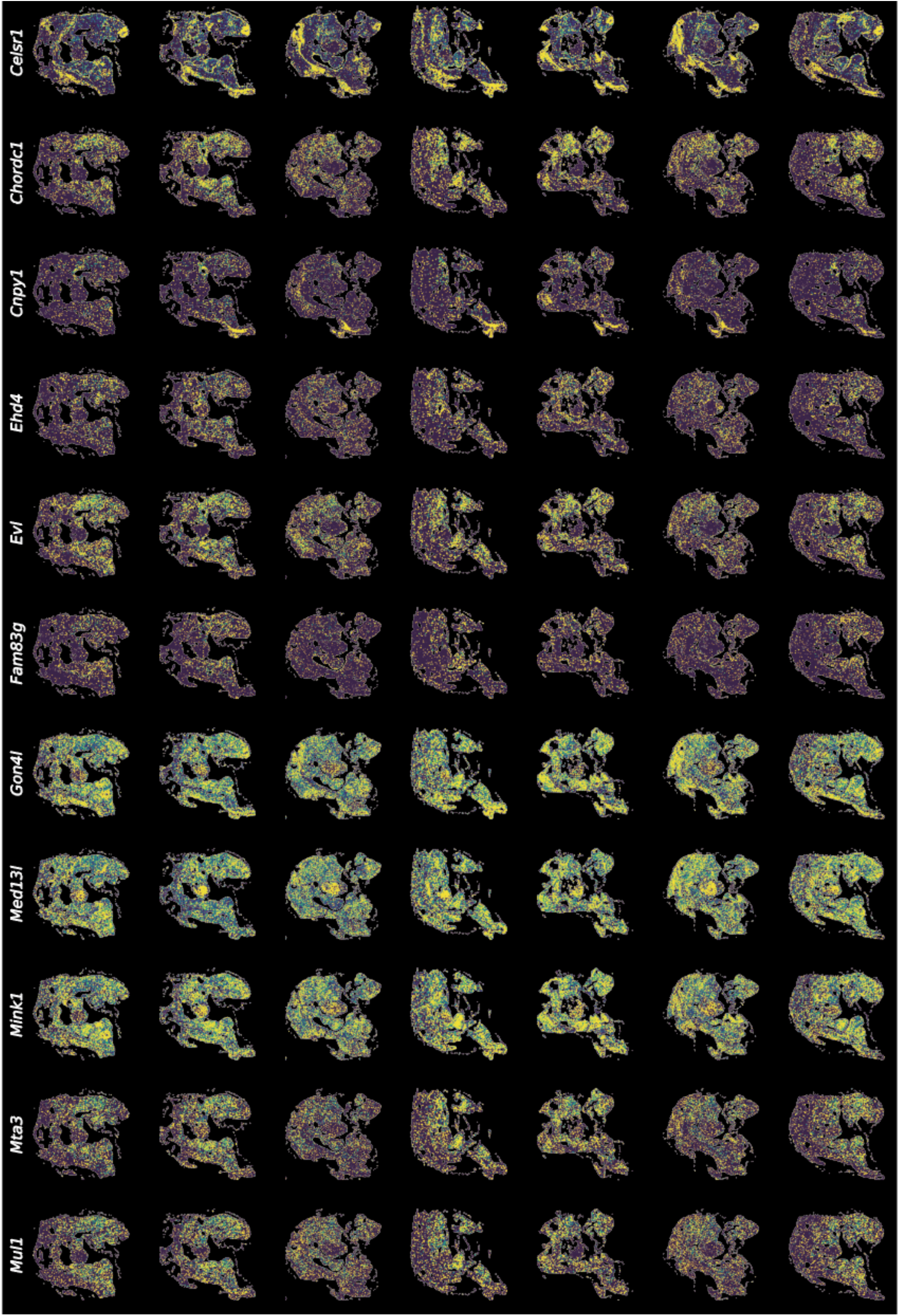
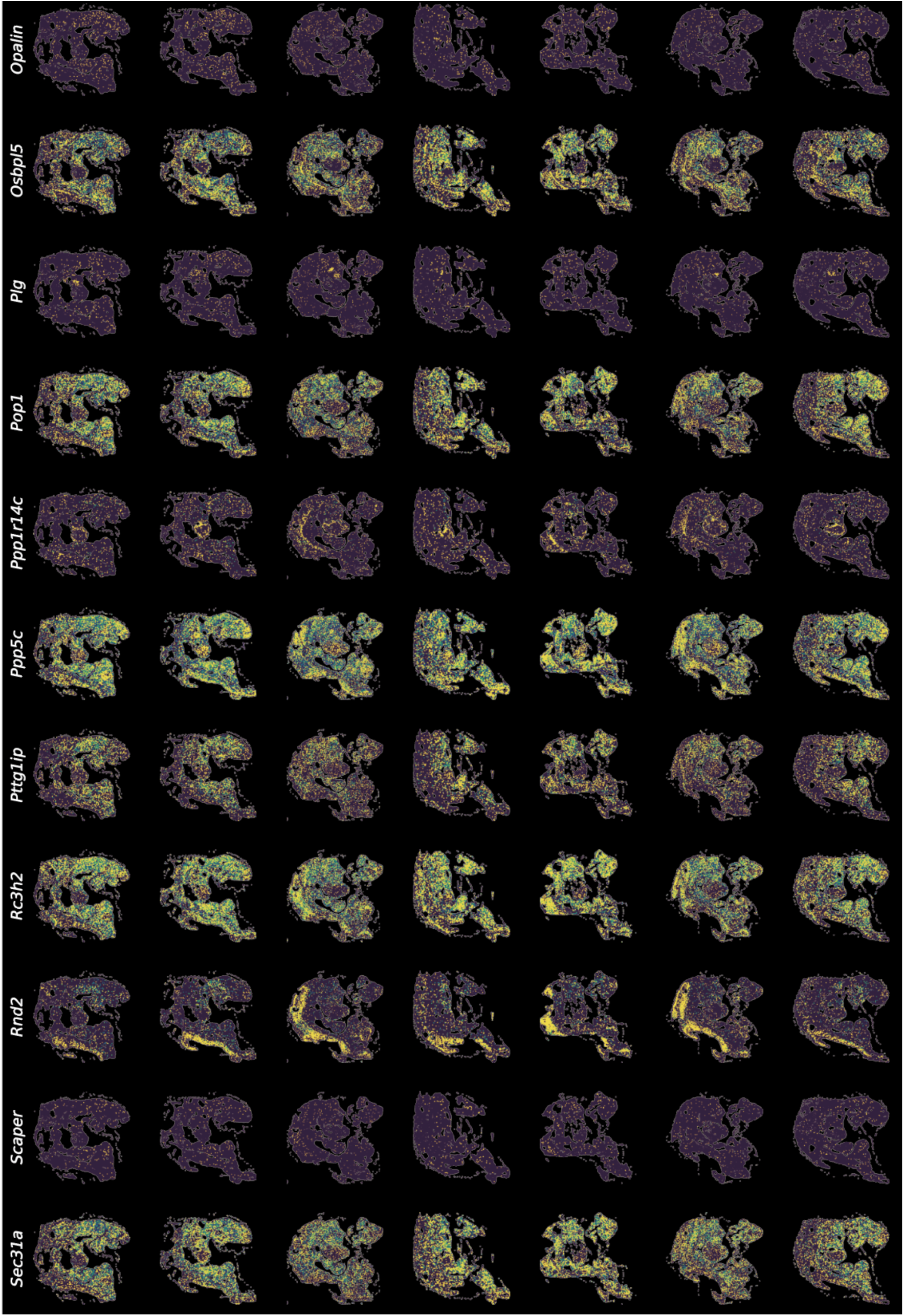
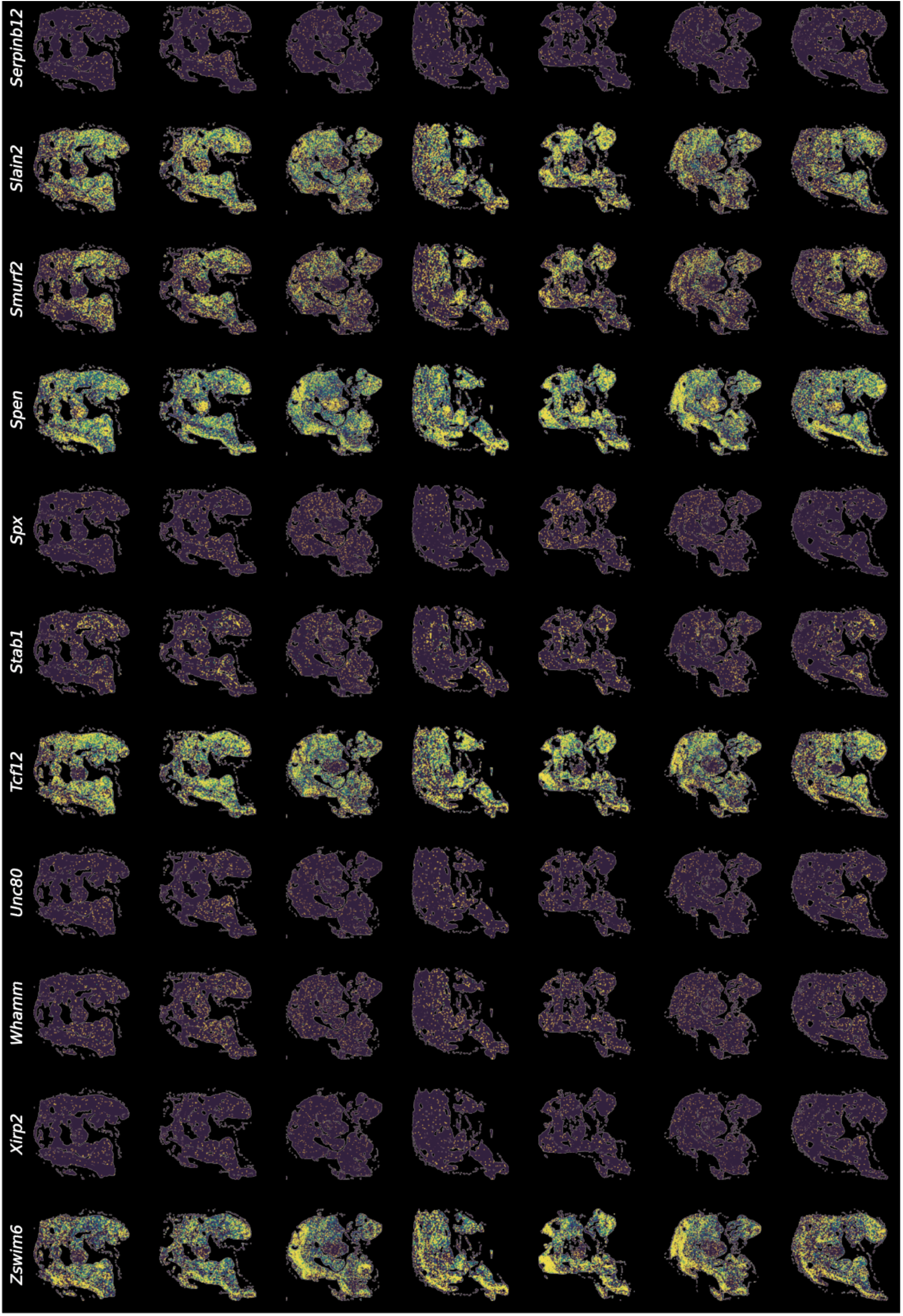
Spatial expression of DNM genes with MERFISH. Spatial expression of the 36 MM genes with damaging DNMs. **a**, Gene expression of marker genes for seven selected cell types (neuron, neural progenitor, pre-epithelial to mesenchymal transition neural Crest progenitor (NC progenitor), neural crest, mesoderm, dorsal root ganglia, and blood), in seven embryonic replicates (regions) 1 (top)-7 (bottom). **b**, Spatial expression pattern of 36 damaging DNM genes. The seven replicates (X axes) of mouse E10.5 showing the gene expression pattern of candidate genes (Y axes). R=Rostral, C=Caudal, V=Ventral, D=Dorsal.

**Extended Data Figure 5.**
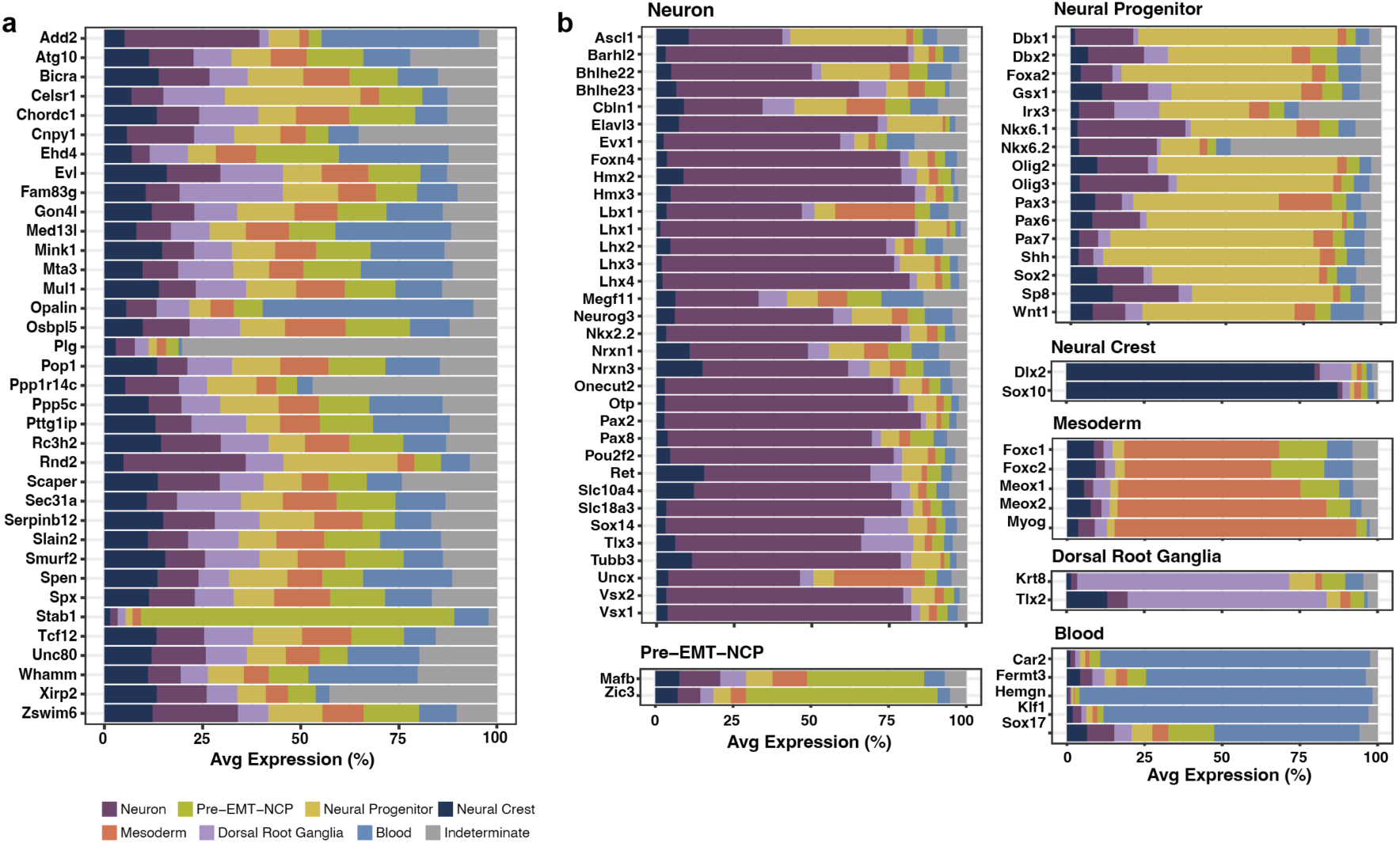
Cell type expression of the DNM genes in MERFISH. **a**, Expression of the 36 damaging DNMs in seven cell types: neuron, neural progenitor, pre-epithelial to mesenchymal transition neural crest progenitor (Pre-EMT-NCP), neural crest, mesoderm, dorsal root ganglia, and blood. Indeterminate refers to the cells that were not specified with the marker genes designed for the seven cell types. **b**, Expression of marker genes used for specifying the cell types in MERFISH. Marker genes are shown within the cell type category which they represent.

**Extended Data Figure 6.**
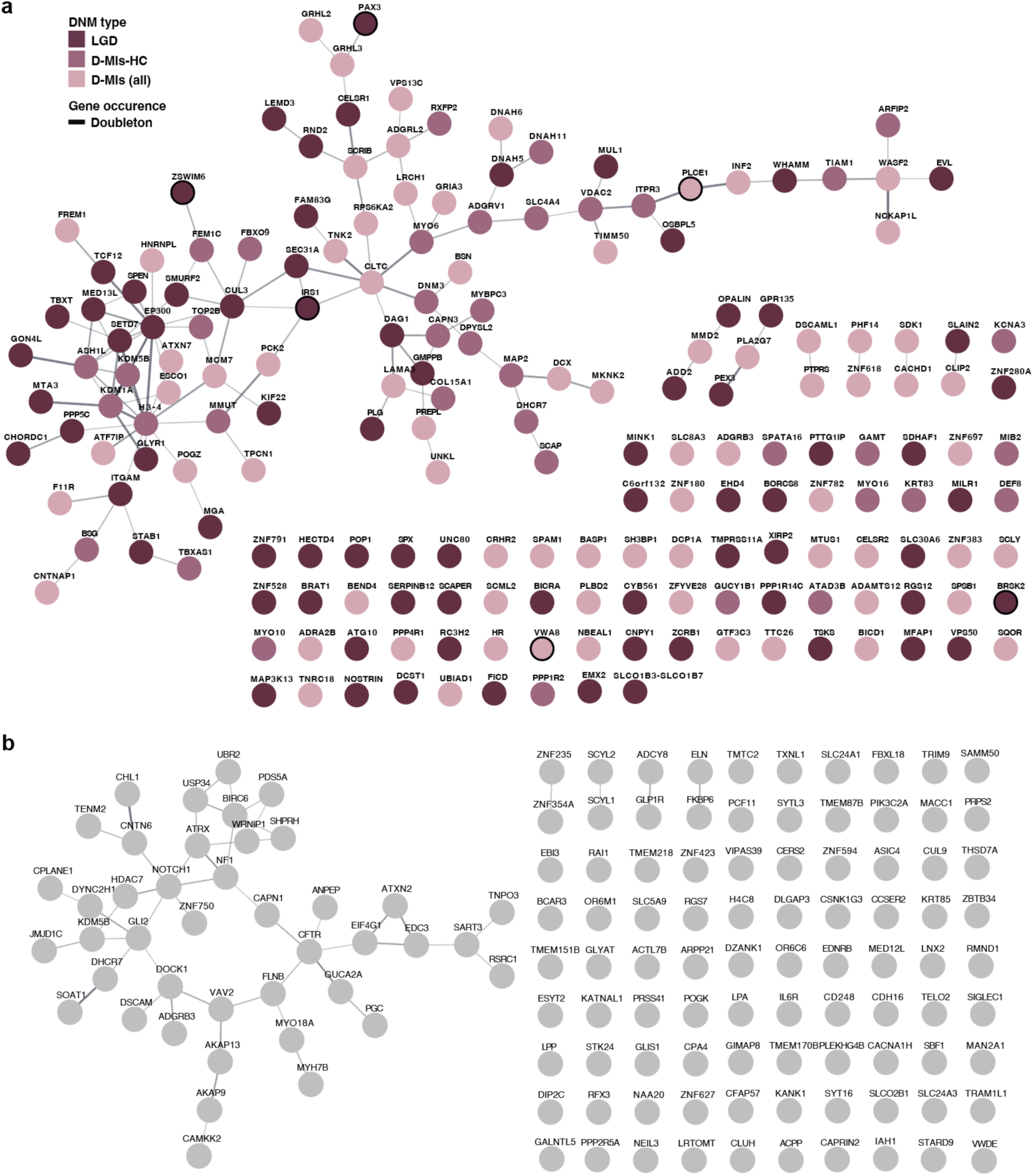
Connections of genes having damaging DNMs of MM and control cohorts. **a-b**, Genes with damaging DNMs (LGD + D-Mis) and their interconnections based on the STRING database, are shown for **(a)** MM and **(b)** control cohorts. Node color: variant functional categories: LGD (purple), D-Mis (light pink), and D-Mis-HC (dark pink). Doubleton genes: black borders. Edge thickness: confidence score of the protein interaction.

**Extended Data Figure 7.**
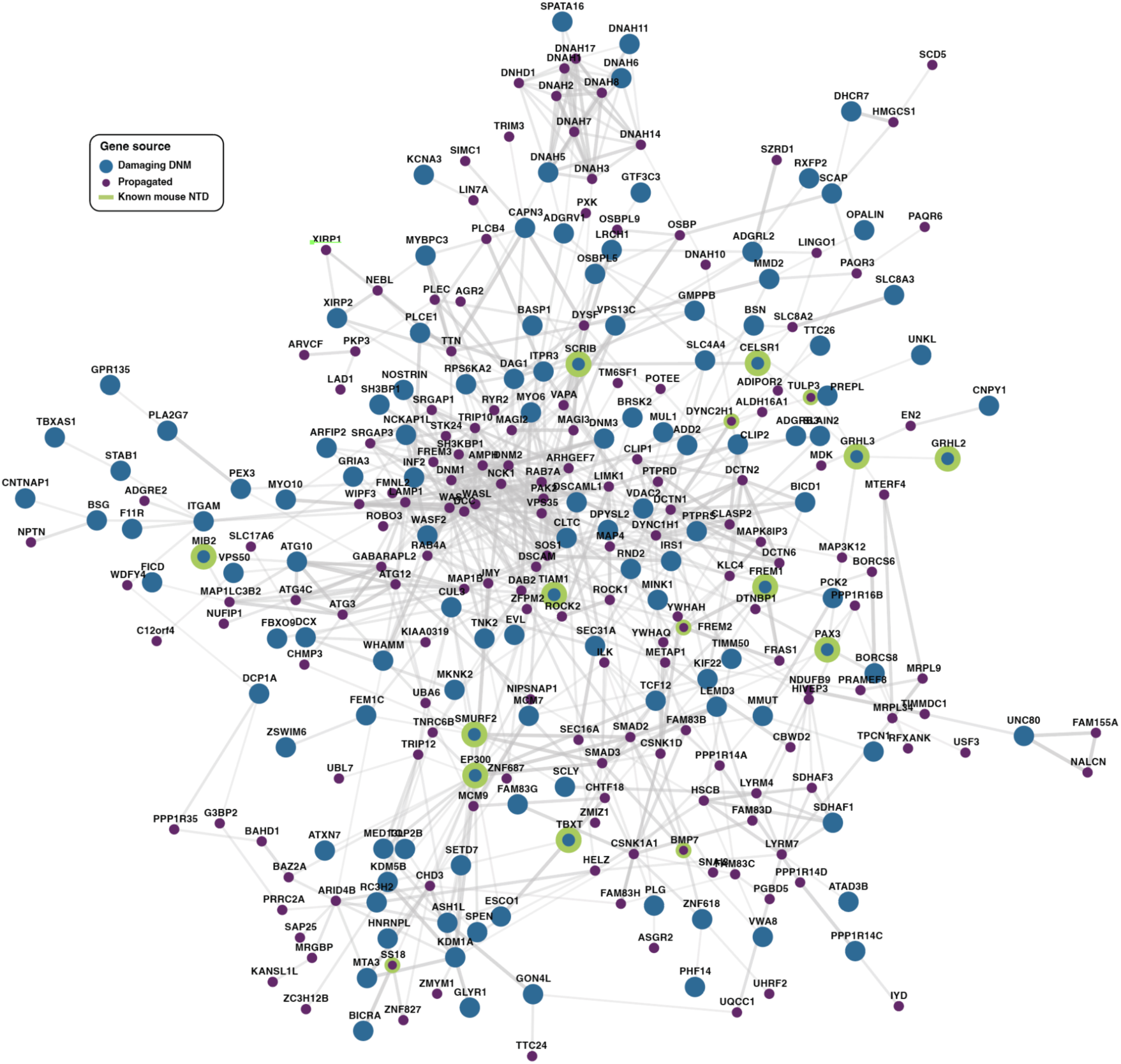
A human ‘Meningomyel-ome’ constructed with damaging DNMs contributing to MM risk. By using the 187 damaging MM DNM genes, a propagated network, Meningomyel-ome, was generated with NetColoc^69^ with a background protein network PCNet^70^, incorporating 439 nodes and 2,447 edges. Big blue circle: damaging DNM genes, Small purple circle: propagated gene, Green border: known mouse NTD genes. The network is visualized with Cytoscape with STRING database.

**Extended Data Figure 8.**
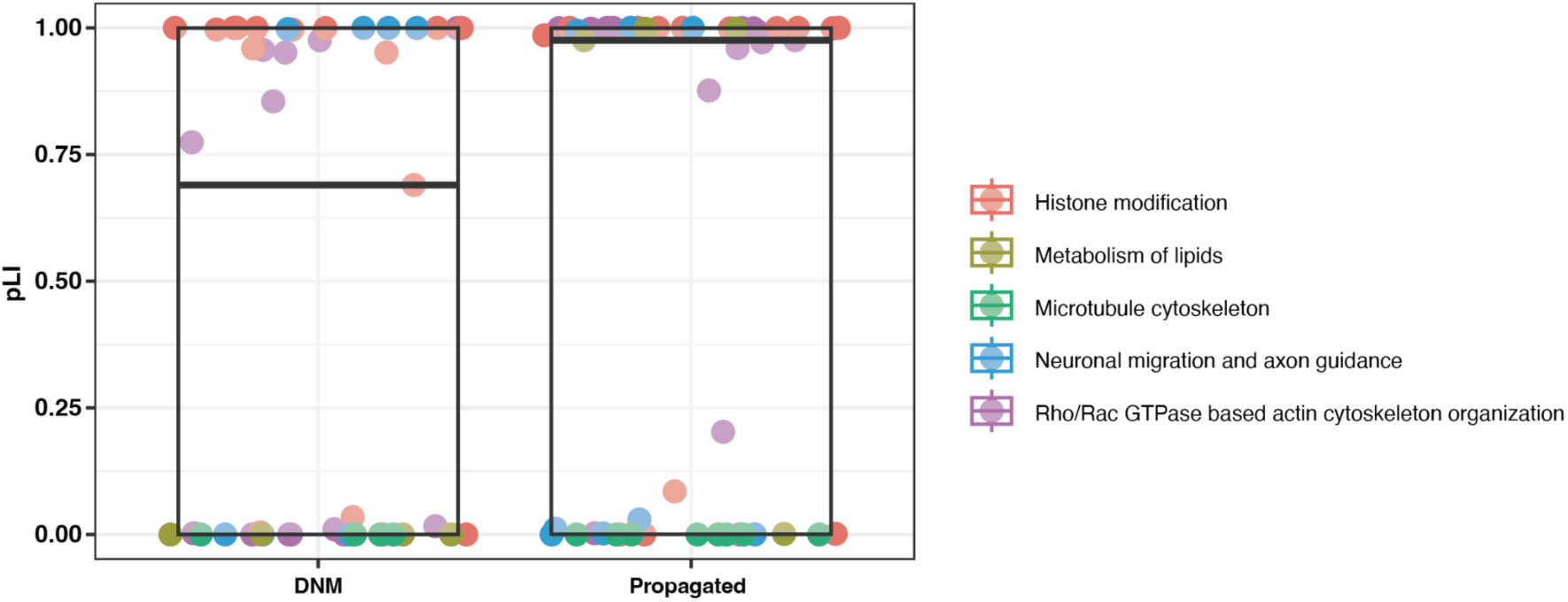
pLI comparison of DNM and propagated genes from the five submodules. The probabilities of being loss of function intolerant (pLI) of genes from the five functional submodules are shown, derived from 46 damaging DNMs and 51 propagated genes. Black thick line: median, Box: 1^st^ and 3^rd^ quantiles.

**Extended Data Figure 9.**
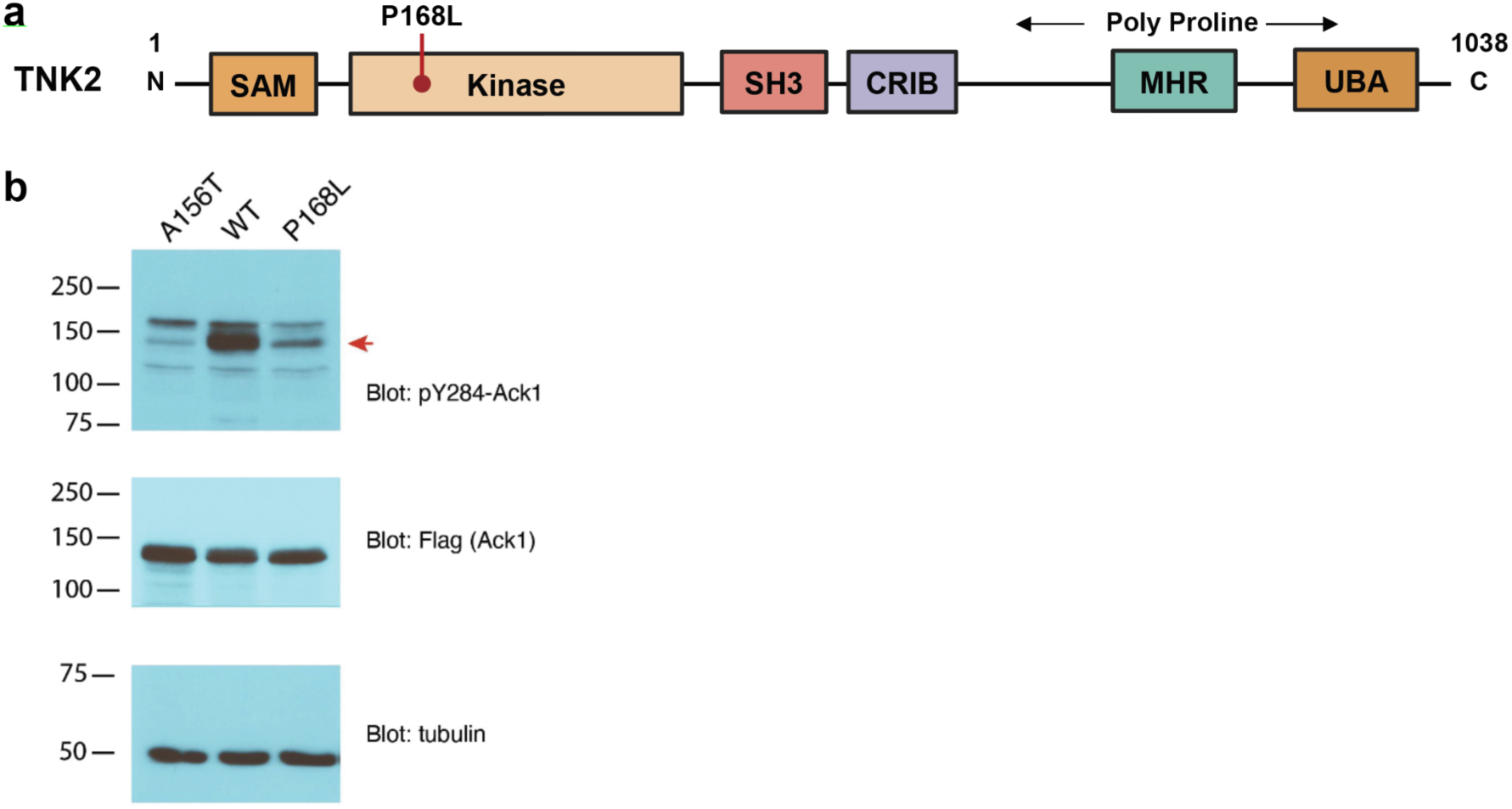
The P168L patient mutation impairs TNK2 activity. **a**, P168L mutation is located within the kinase domain. TNK2 contains sterile alpha motif (SAM), Src homology 3 (SH3), CDS42, and RAC-interactive binding (CRIB), Mig6 homology region (MHR), and ubiquitin-associated domain (UBA). **b**, Blots for the A156T kinase dead, wild-type, and the patient mutation P168L. Lysates were probed with pY284-Ack1 (top), Ack1-flag (middle), and gamma-tubulin (bottom).

**Extended Data Figure 10.**
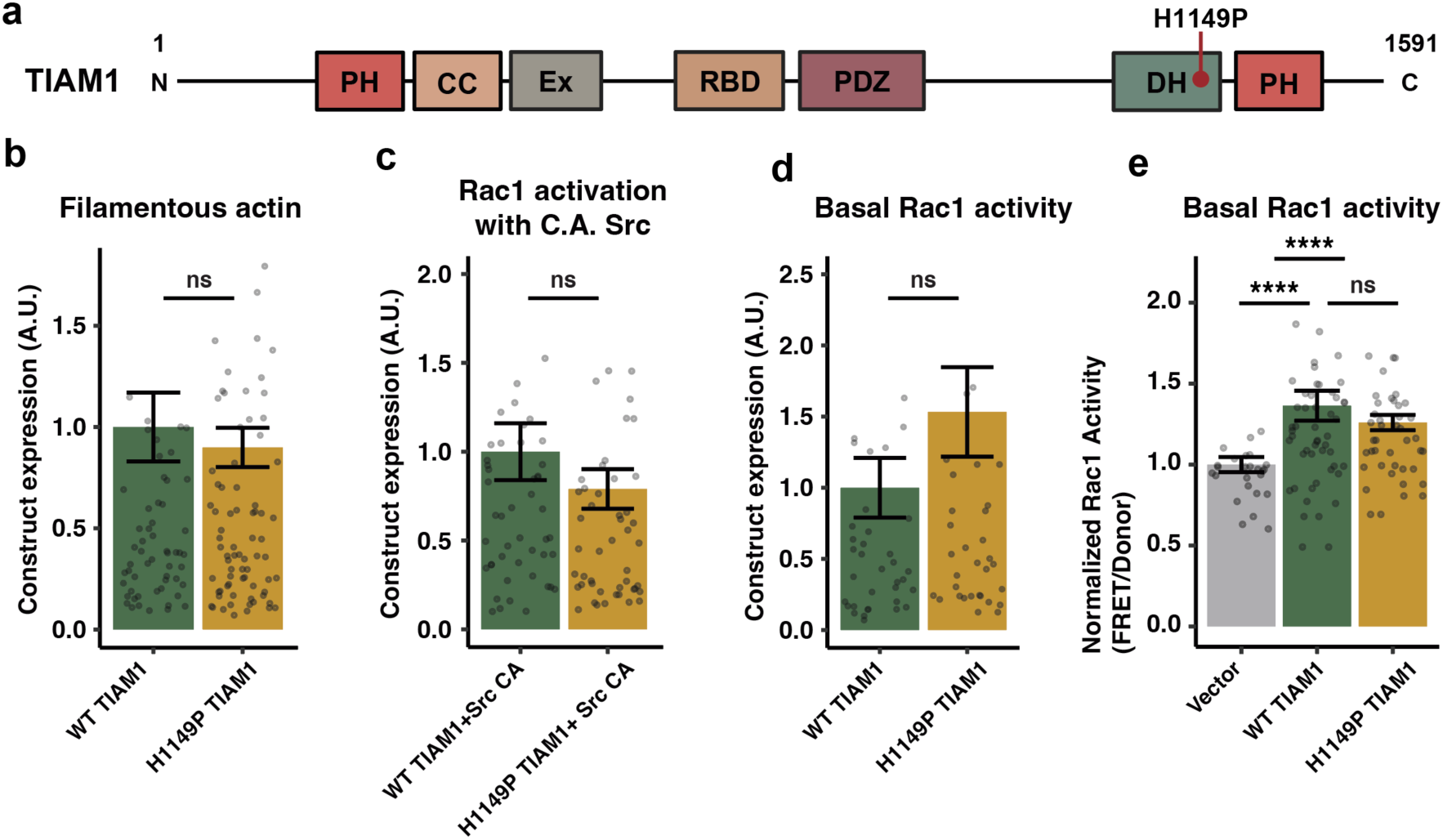
The H1149P patient mutation impairs TIAM1 activity. **a**, H1149P mutation is located within the Dbl homology (DH) domain responsible for GEF activity. TIAM1 contains an N-terminal pleckstrin homology (PH), coiled-coiled (CC), extension (Ex), RAS binding (RBD), PDZ, Dbl-homology (DH) and PH domains, with the patient mutation falling within the DH domain. **b**, Construct expression H1149P is equivalent to wildtype in Phalloidin quantification. **c-d**, Construct expression H1149P in (**c**) basal and (**d**) constitutive active (C.A.) Src Rac1 Förster resonance energy transfer (FRET) is equivalent to wildtype. **e**, Protein expression H1149P in C.A. Src Rac1 FRET is equivalent to wildtype. Bar: mean, Errorbar: standard deviation of mean (SEM). Kruskal-Wallis followed by a pairwise Wilcoxon test, *P* value adjusted with Bonferroni. Data shown with Hampel filter.

**Extended Data Figure 11.**
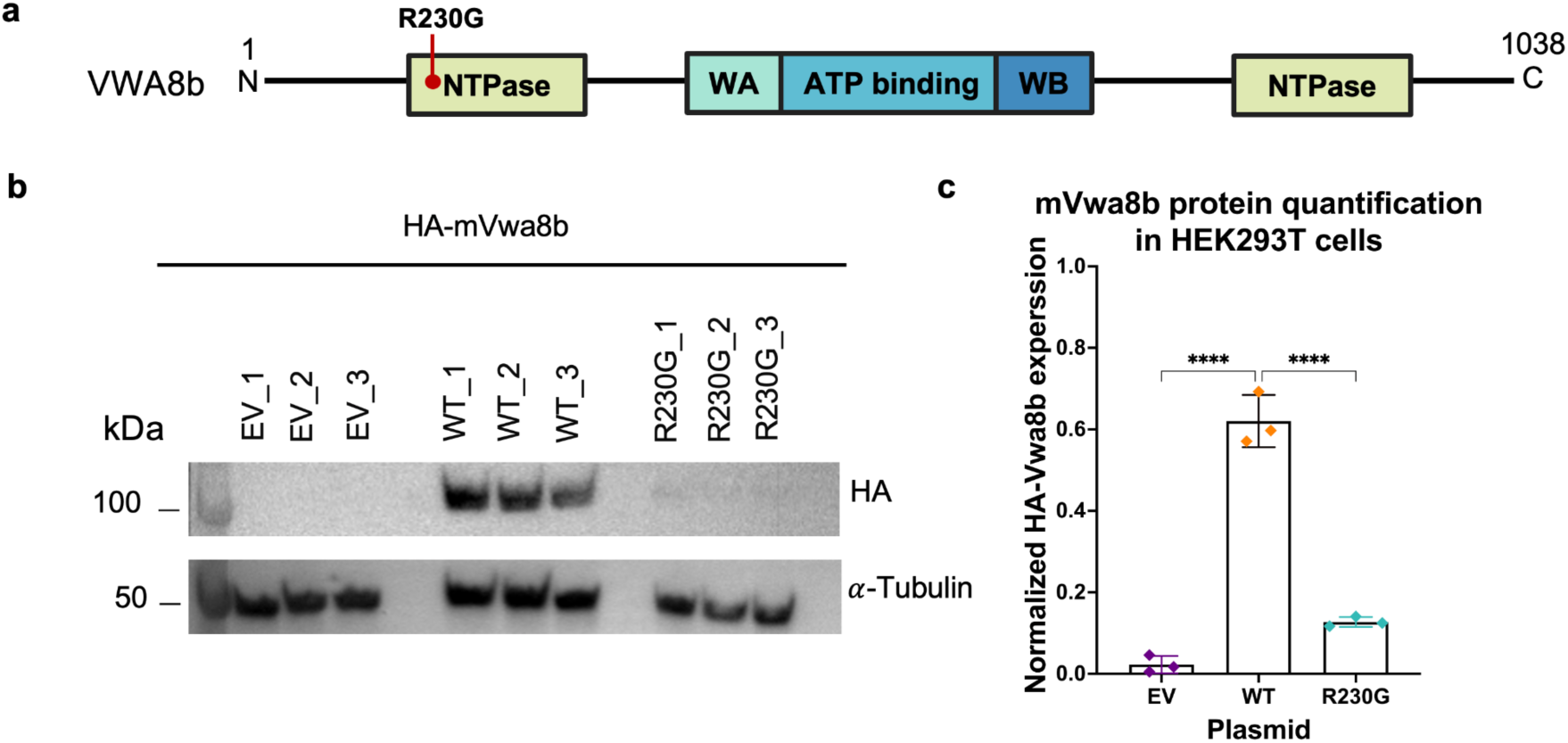
*VWA8b* patient mutation R230G significantly reduces protein expression levels. **a,** Schematic of protein domains for VWA8b consisting of NTPase, Walker A (WA), ATP binding, and Walker B (WB) domain and patient mutation R230G in the NTPase domain. **b,** Western blot for HA-tagged mVwa8b empty vector (EV), wildtype and p.R230G protein overexpressed in HEK293T cells and stained for HA and alpha tubulin as loading control**. c,** Quantification of HA band intensity from panel **b** normalized to loading control. Bar: mean, Errorbar: standard deviation of mean (SEM). one-way ANOVA ****: *P* value < 0.0001.

**Extended Data Figure 12.**
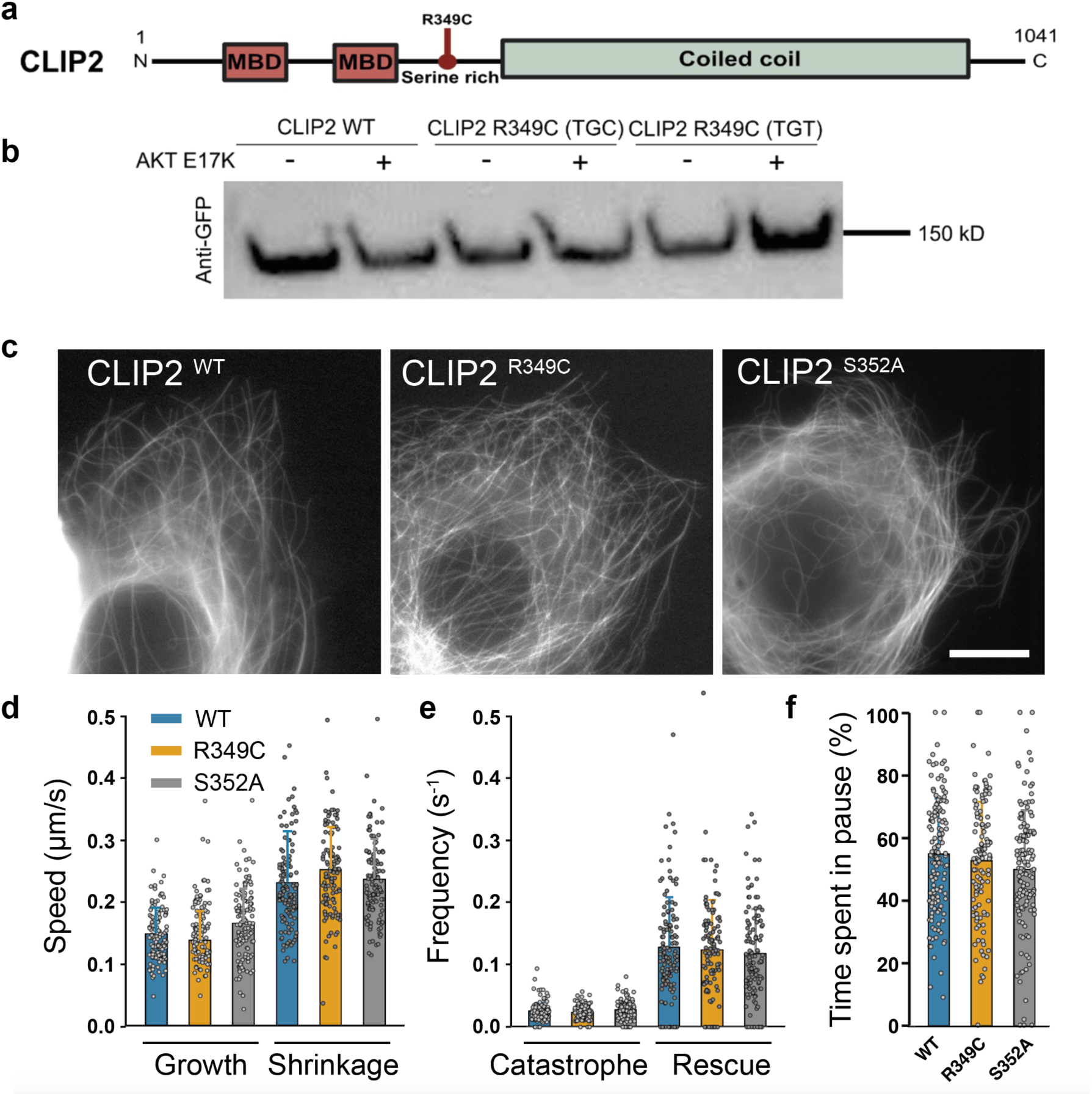
The R349C patient mutation in *CLIP2* has no observable effect on microtubule dynamics. **a,** Domains in CLIP2 protein, microtubule associated domain (MBD), serine rich region containing the R349C patient mutation, and coiled coil domain. **b,** GFP fused CLIP2 expression in HEK293T cells for wildtype (WT) and two synonymous codons (TGC and TGT both encoding for CYS) for R349C patient mutation. No notable difference in the level of expression was detected between the WT and the mutants. Experiments included co-transfection with constitutively active AKT E17K, a presumed upstream kinase of nearby S352. **c-f,** While at a low expression level in HuH7 cells, fluorescent CLIP2 is restricted to growing microtubule plus ends as expected (not shown), (**c**) its mild overexpression highlights the whole microtubule bodies without difference between WT, R349C (TGC) and a putative phosphorylation site, S352A (Scale bar 10µm). This condition allowed the measurement of a complete panel of microtubule dynamic instability parameters: (**d**) growth and shrinkage speed, (**e**) catastrophe and rescue (R) frequencies (**f**) the time spent in pause. There was no statistical difference between CLIP2 WT and the mutants. The values were measured from at least 125 microtubules (25 cells) per condition. n = 3 independent experiments. Bar: mean, Point: data for each microtubule, Errorbar: standard deviation.

**Extended Data Figure 13.**
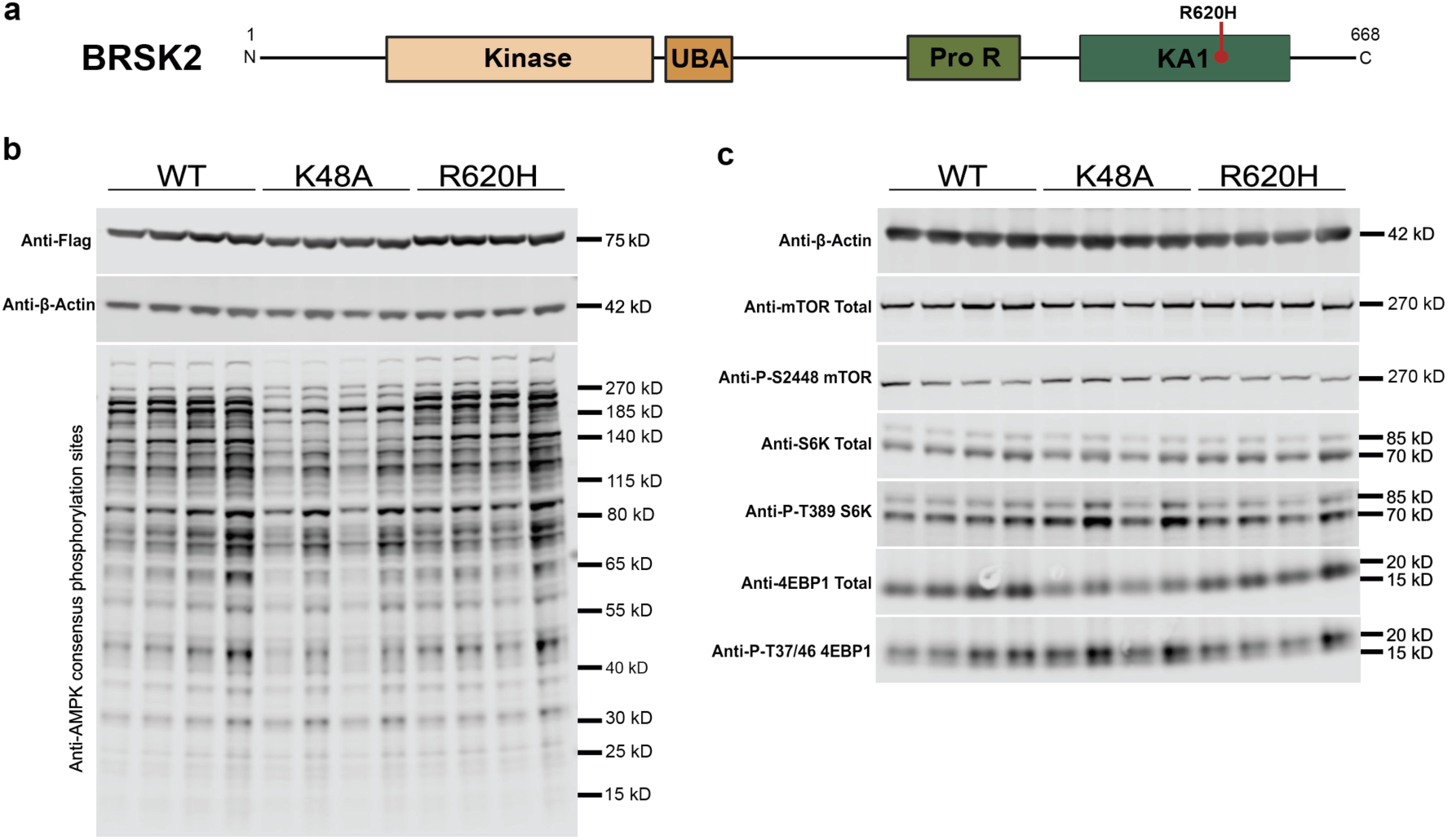
*BRSK2* patient mutation R620H shows no notable defects in kinase activity across known downstream phosphorylation targets. **a,** R620H mutation is located within the kinase associated 1 domain (KA1). BRSK2 also contains a kinase domain, Ubiquitin-associated (UBA), Proline rich region (Pro R). **b,** Western blot showing phosphorylation of AMPK substrate motifs observed across wildtype (WT), kinase dead mutant control (K48A) and patient mutation (R620H) Flag-tagged BRSK2 when expressed in HEK293T cells. K48A has an average reduced intensity that is not observed in R620H. **c,** Western Blots of total and phosphorylated mTOR, S6K and 4EBP1 substrates showing no notable differences in phosphorylation for patient mutation R620H.

**Extended Data Figure 14.**
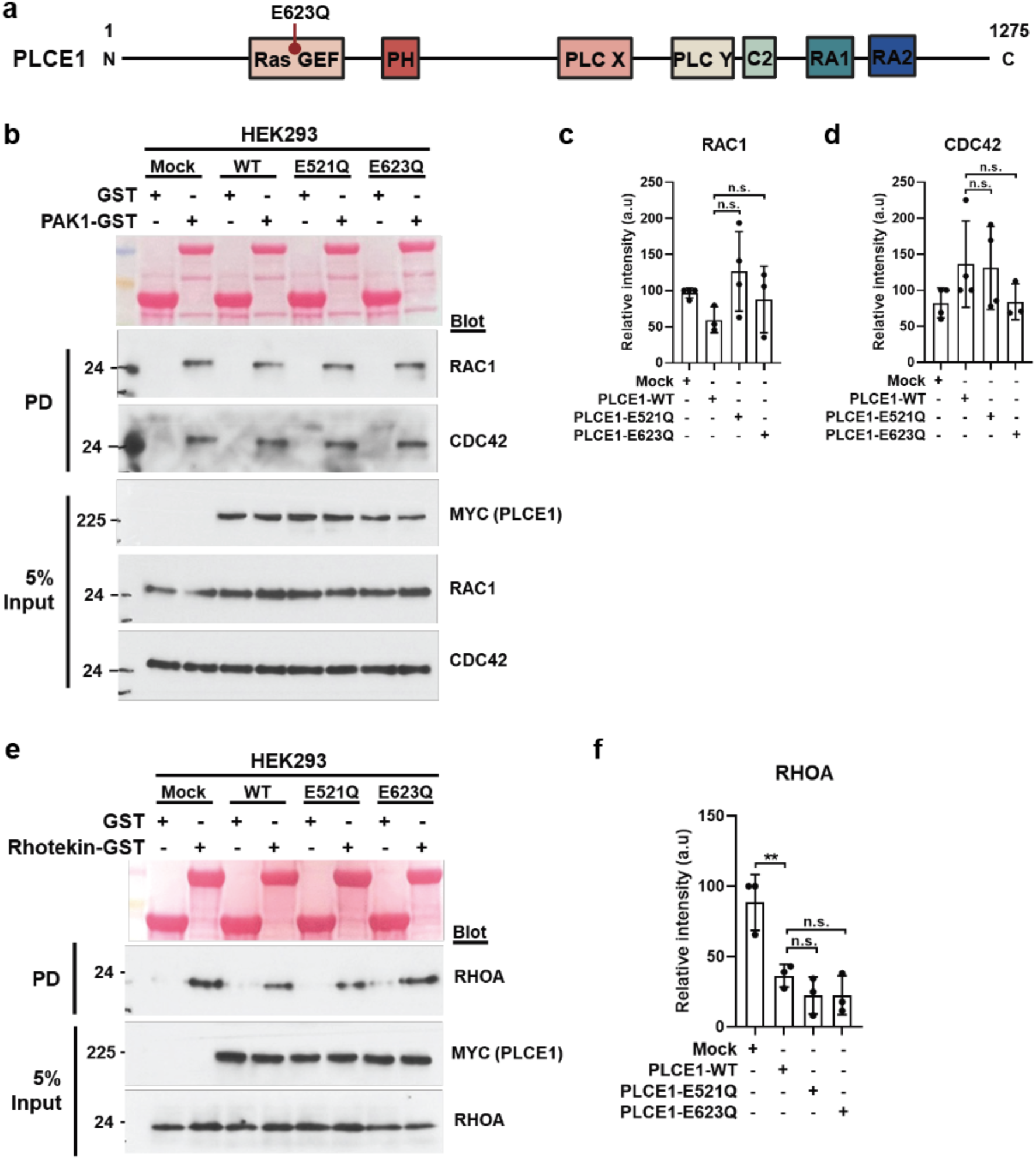
*PLCE1* patient mutation E623Q does not lead to diminished Rho GTPase activity. **a,** Schematic of PLCE1 protein with domains annotated. Patient E623Q mutation is located in the Ras GEF domain. PLCE1 contains a Guanine nucleotide exchange factor for Ras-like small GTPases (RAS GEF), Pleckstrin Homology (PH), Phospholipase C catalytic domain X (PLCX), Phospholipase C catalytic domain Y (PLCY), Protein Kinase C conserved region 2 (C2), RAS association domain 1 (RA1), and RAS association domain 2 (RA2). **b,** Active GTP-bound forms of RAC1 and CDC42 precipitated from HEK293 expressing Myc-tagged PLCE1 using a GST-PAK1 pulldown assay (n=4). Five percent input represents the controls for equal loading. Compared to mock cells, overexpression of wild-type (WT) and variant forms of PLCE1 exhibited no significant difference in RAC1 and CDC42 activity. **c**-**d**, Quantifications of (**c**) RAC1 and (b) CDC42. **e**, Active GTP-bound form of RHOA precipitated from HEK293 expressing Myc-tagged PLCE1 using a GST-rhotekin pulldown assay. Overexpression of WT PLCE1 resulted in a substantial decrease in relative RHOA activity compared with mock cells. Compared to WT, cells transfected with variant forms of PLCE1 exhibited no significant differences in GTP-bound RHOA. **f**, Quantification of RHOA (n=4). Data were analyzed by one-way ANOVA with post hoc Bonferroni correction. Error bars: standard deviation for greater than 3 independent experiments. PD; pulldown, ** P < 0.01, n.s. not significant.

**Extended Data Fig. 15.**
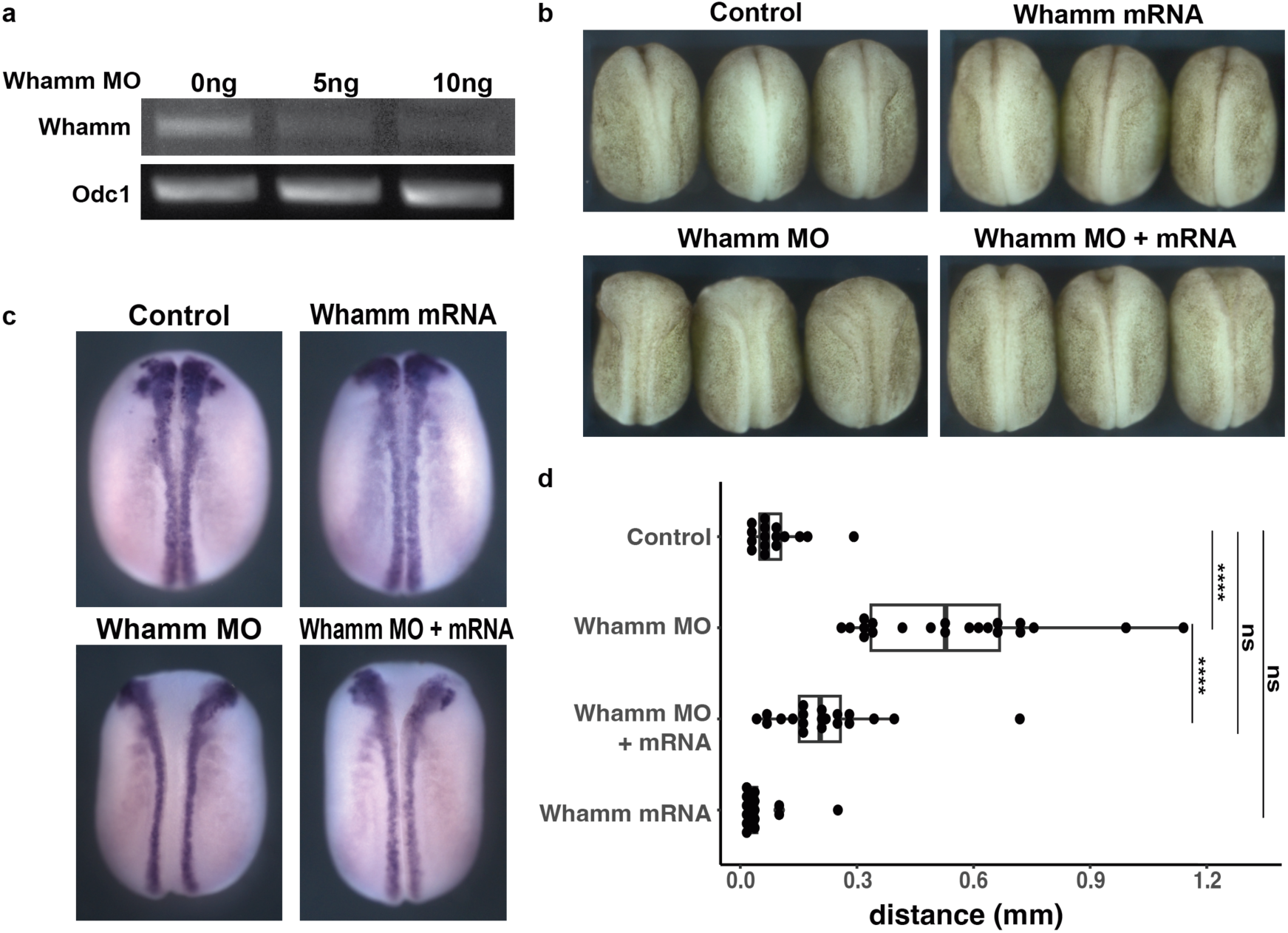
Validation of Whamm morpholino antisense oligonucleotide (MO). **a,** RT-PCR validates the effective disruption of splicing following injection of Whamm MO. **b-d**, The neural tube closure defect phenotype induced by Whamm MO (10 ng) was rescued through the injection of Whamm mRNA (700 pg). Embryos injected only with mRNA showed no significant phenotype. **b**, Dorsal views of Xenopus embryos at Stage 19 **c**, Embryos subjected to in situ hybridization against Pax3 to visualize the neural folds **d**, Graph depicting the quantitative analysis of the distance between neural folds. P-values by one-way ANOVA with post-hoc honestly significant difference (HSD) test. ****: *P* value < 0.0001, ns: not significant.

**Extended Data Figure 16.**
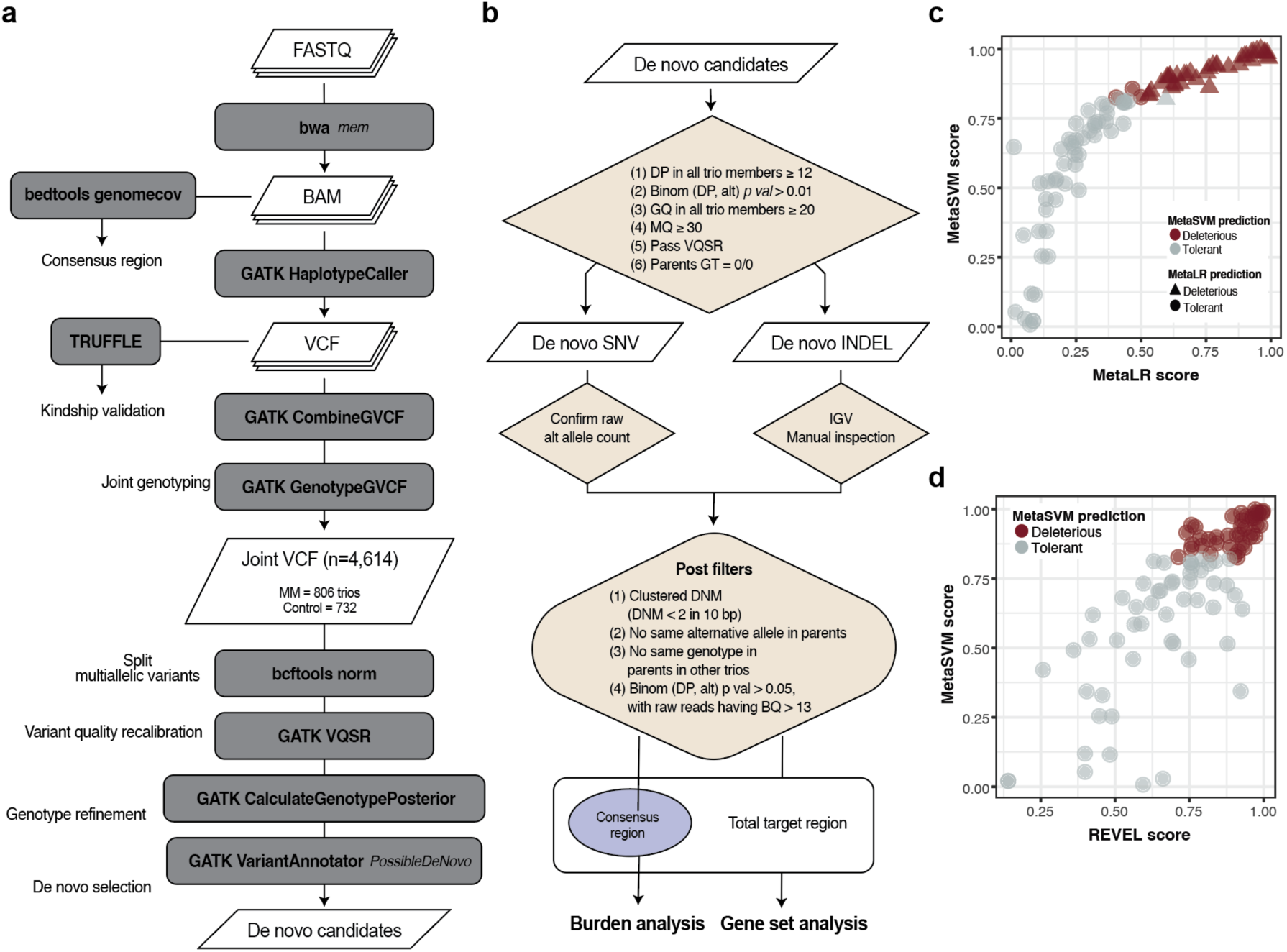
Pipeline of detecting high-confidence de novo mutation (DNM)s and annotation of D-Mis-HC. **a**, Schematic overview of DNM detection. The raw coverages of aligned bam files were utilized in consensus region generation (see Methods). Germline variants were utilized to test the integrity of the trios with TRUFFLE. GATK genotype refinement workflow was utilized for the DNM detection **b**, The candidate de novo SNVs and Indels were applied with a series of qualitative and quantitative filters. DP: depth, Binom: binomial test, alt: count of reads with alternative alleles, GQ: genotype quality, MQ: mapping quality, VQSR: variant quality score recalibration, GT: genotype. **c-d**, Pathogenicity prediction of D-Mis-HC variants with MetaLR and REVEL. The pathogenicity of 116 D-Mis DNMs predicted by MetaSVM are shown with the prediction results from MetaLR and REVEL. (**c**) MetaLR deleterious (score > 0.5) is shown in triangle and (**d**) mutations with REVEL score higher than 0.75 are predicted as deleterious (right).

## Extended Data Table

**Extended Data Table 1.**
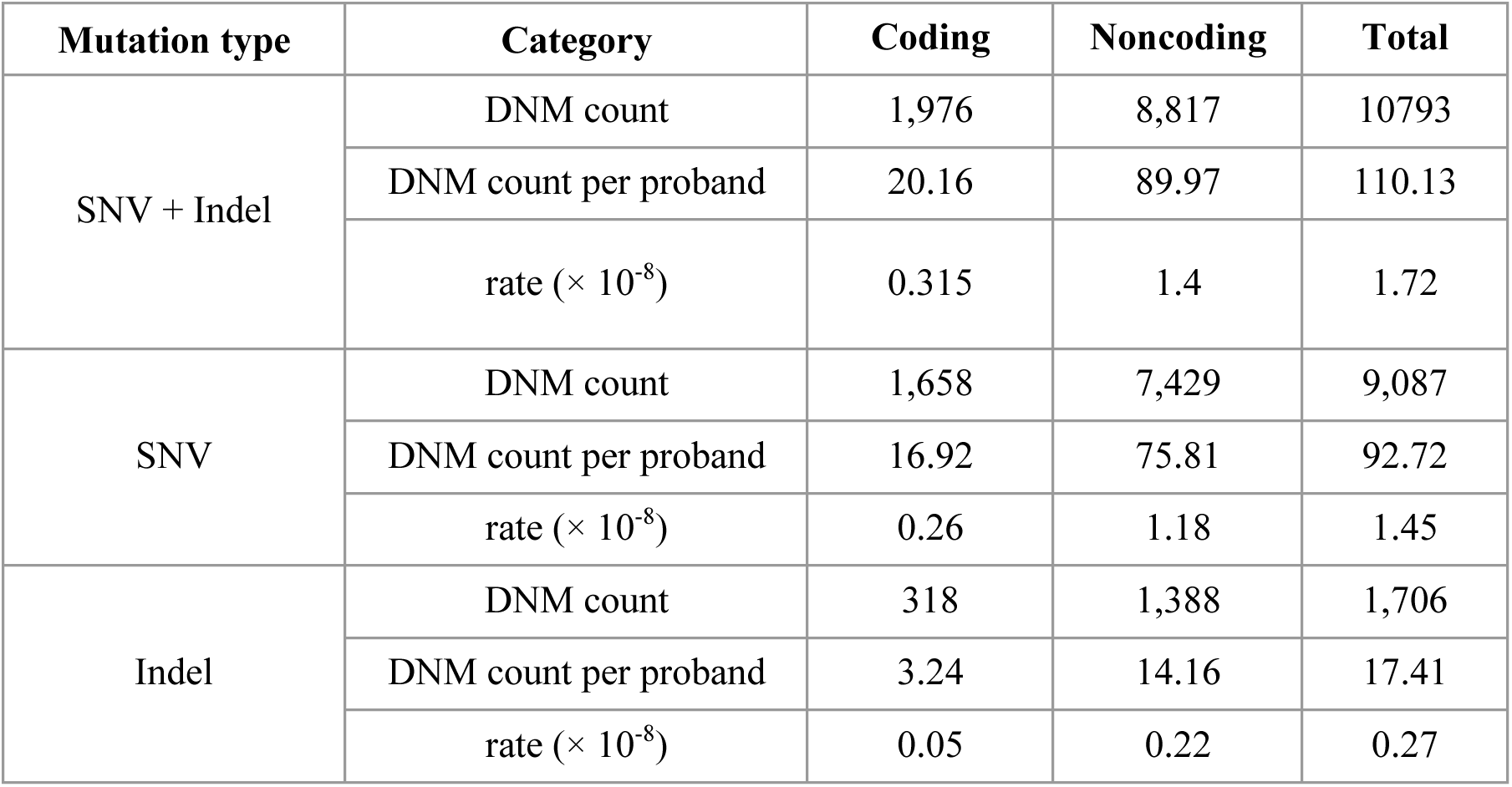
De novo SNV and Indel rates (×10^-8^) of WGS in coding and noncoding regions. Rates were calculated with 99 trios could be analyzed after removal of four kinship failed trios.

| Mutation type | Category | Coding | Noncoding | Total |
| --- | --- | --- | --- | --- |
| SNV + Indel | DNM count | 1,976 | 8,817 | 10793 |
|  | DNM count per proband | 20.16 | 89.97 | 110.13 |
| | rate ($\times 10^{-8}$) | 0.315 | 1.4 | 1.72 |
| SNV | DNM count | 1,658 | 7,429 | 9,087 |
|  | DNM count per proband | 16.92 | 75.81 | 92.72 |
| | rate ($\times 10^{-8}$) | 0.26 | 1.18 | 1.45 |
| Indel | DNM count | 318 | 1,388 | 1,706 |
|  | DNM count per proband | 3.24 | 14.16 | 17.41 |
| | rate ($\times 10^{-8}$) | 0.05 | 0.22 | 0.27 |

**Extended Data Table 2.** de novo large CNV (>100kb) in WGS, analyzed with 99 trios.

| Family | Chromosome | Start position | End position | Length | Variant type | Cytoband |
| --- | --- | --- | --- | --- | --- | --- |
| 7646 | chr22 | 20,400,001 | 21,100,001 | -700,000 | Deletion | q11.21 |
| 7647 | chr15 | 72,700,001 | 75,800,001 | -3,100,000 | Deletion | q24.2-q24.1 |
| 7662 | chr15 | 30,600,001 | 32,100,001 | 1,500,000 | Duplication | q13.3-q13.2 |
| 8469 | chr21 | 13,900,001 | 43,000,001 | 28,900,000 | Duplication | q22.11-q22.12 |

**Extended Data Table 3.** Pathway enrichment of the damaging DNMs in MM. 187 damaging DNMs (LGD + D-Mis) were tested for enrichment in known functional biological pathways in Gene ontology (GO) Biological Process, Reactome, and KEGG pathways. Pathways that were significant with Bonferroni step-down adjusted *P* value < 0.05 are shown.

| Term | Ontology Source | ID | *P* Value<br>(Bonferroni<br>step-down<br>adjusted) | %<br>Associated<br>Genes | Nr.<br>Genes | Associated Genes |
| --- | --- | --- | --- | --- | --- | --- |
| **Morphogenesis of**<br>**a polarized**<br>**epithelium** | GO Biological<br>Process | GO:0001738 | 0.00001 | 8.41 | 9.00 | BRSK2, CELSR1,<br>CELSR2, GRHL3,<br>LAMA3, SCRIB,<br>SH3BP1, SMURF2,<br>TIAM1 |
| **Signaling by**<br>**NTRKs** | Reactome | R-HSA:166520 | 0.00031 | 5.97 | 8.00 | CLTC, DNM3,<br>EP300, IRS1,<br>PTPRS, RPS6KA2,<br>TCF12, TIAM1 |
| **Establishment of**<br>**planar polarity** | GO Biological<br>Process | GO:0001736 | 0.00070 | 7.41 | 6.00 | BRSK2, CELSR1,<br>CELSR2, GRHL3,<br>SMURF2, TIAM1 |
| **Wnt signaling**<br>**pathway, planar**<br>**cell polarity**<br>**pathway** | GO Biological<br>Process | GO:0060071 | 0.00100 | 8.62 | 5.00 | CELSR1, CELSR2,<br>GRHL3, SMURF2,<br>TIAM1 |
| **L1CAM**<br>**interactions** | Reactome | R-HSA:373760 | 0.00220 | 5.04 | 6.00 | CLTC, CNTNAP1,<br>DCX, DNM3,<br>DPYSL2, RPS6KA2 |
| **Signaling by**<br>**NTRK1 (TRKA)** | Reactome | R-HSA:187037 | 0.00276 | 5.22 | 6.00 | CLTC, DNM3,<br>EP300, IRS1,<br>RPS6KA2, TCF12 |
| **Neural tube**<br>**closure** | GO Biological<br>Process | GO:0001843 | 0.00323 | 4.90 | 5.00 | CELSR1, GRHL2,<br>GRHL3, SCRIB,<br>TBXT |

## Supplementary Table

**Supplementary Table 1.**
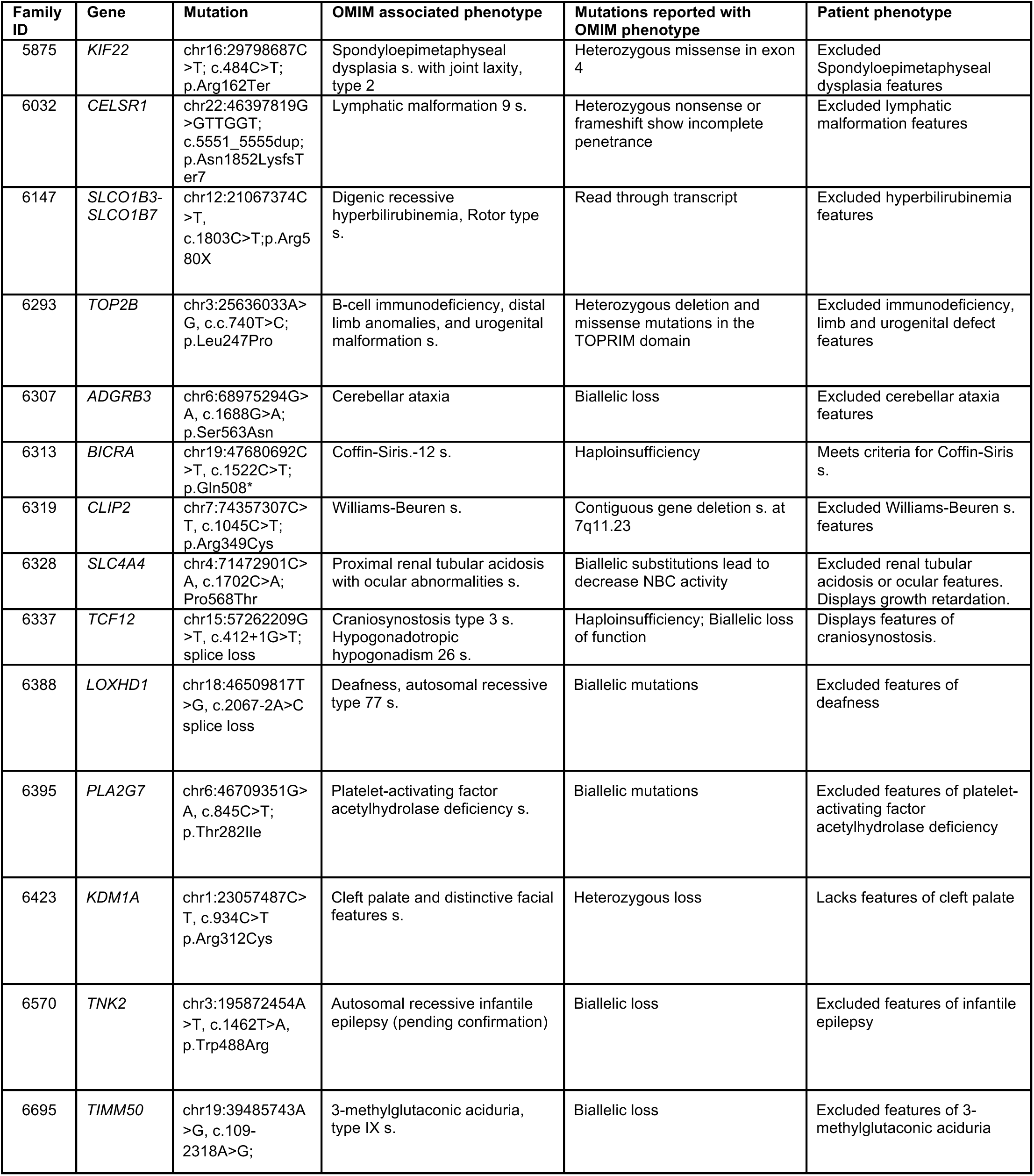
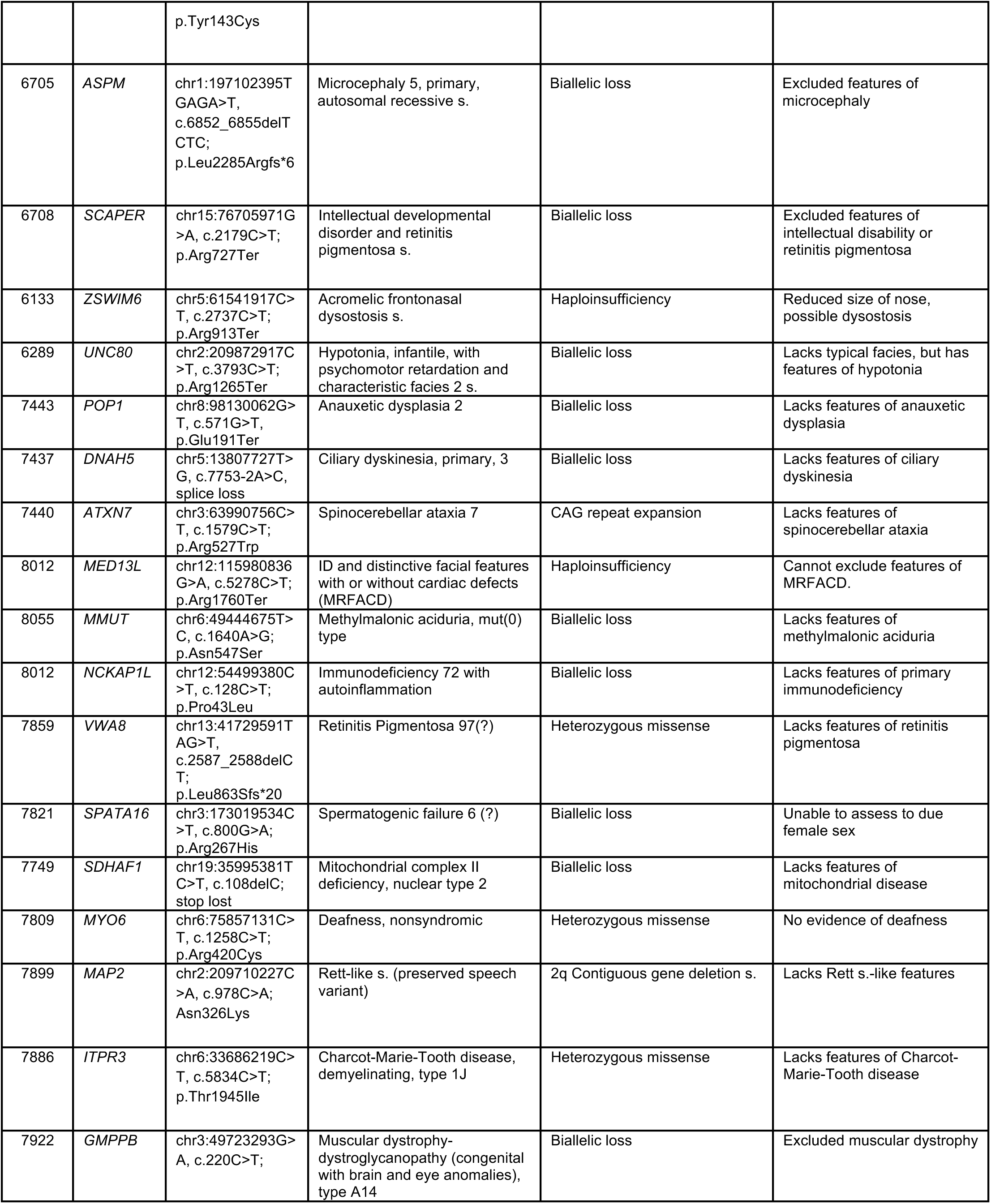
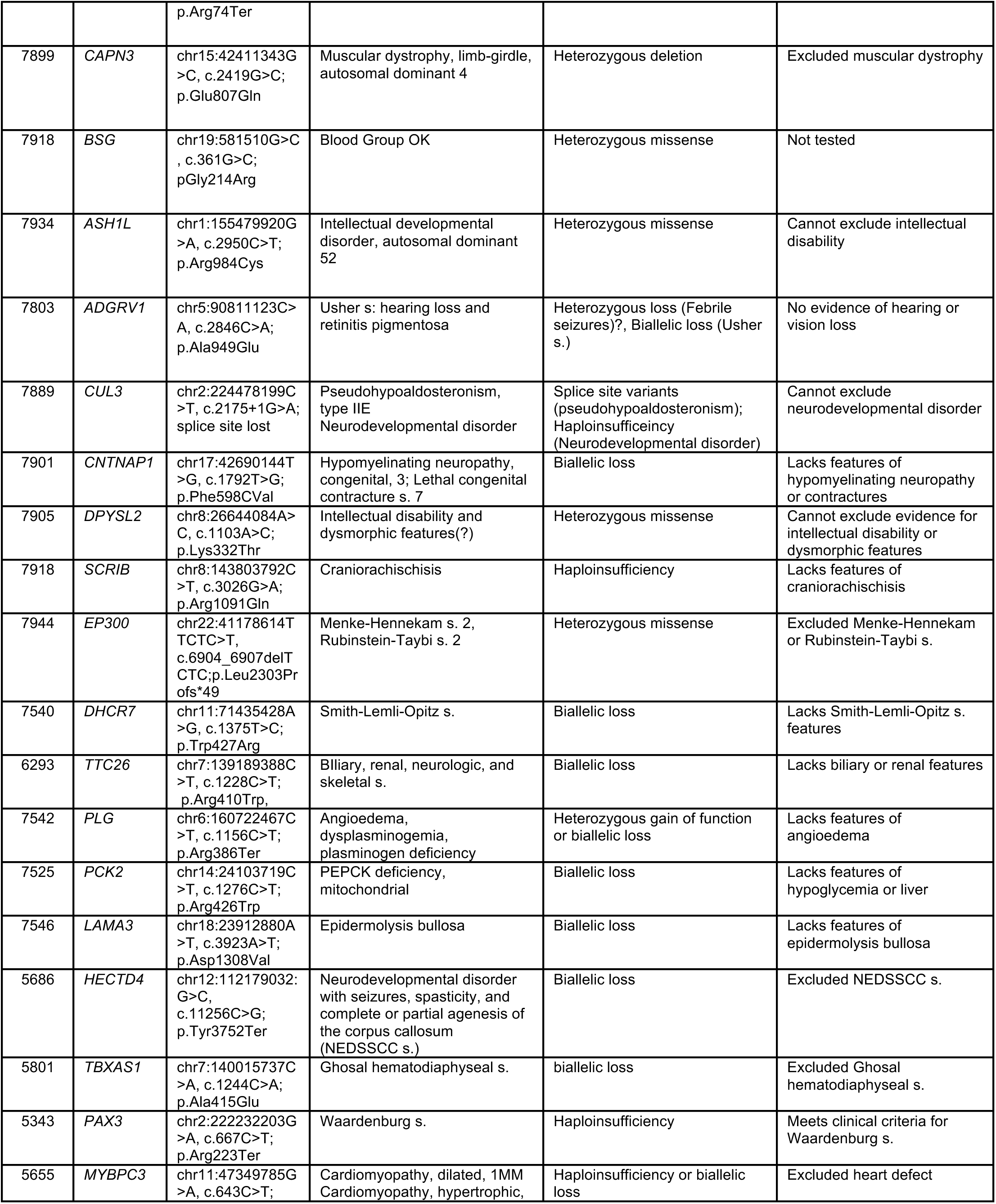
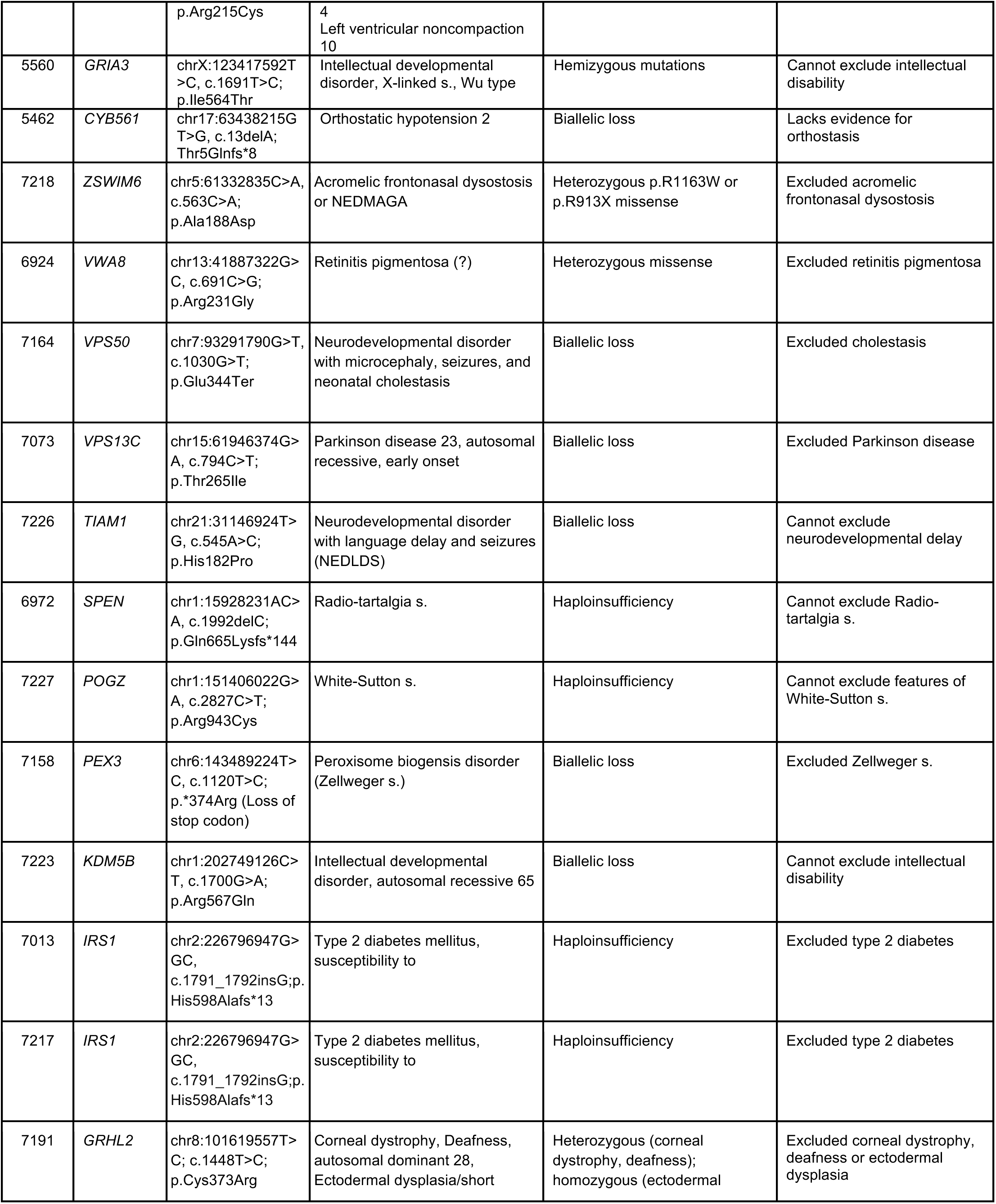
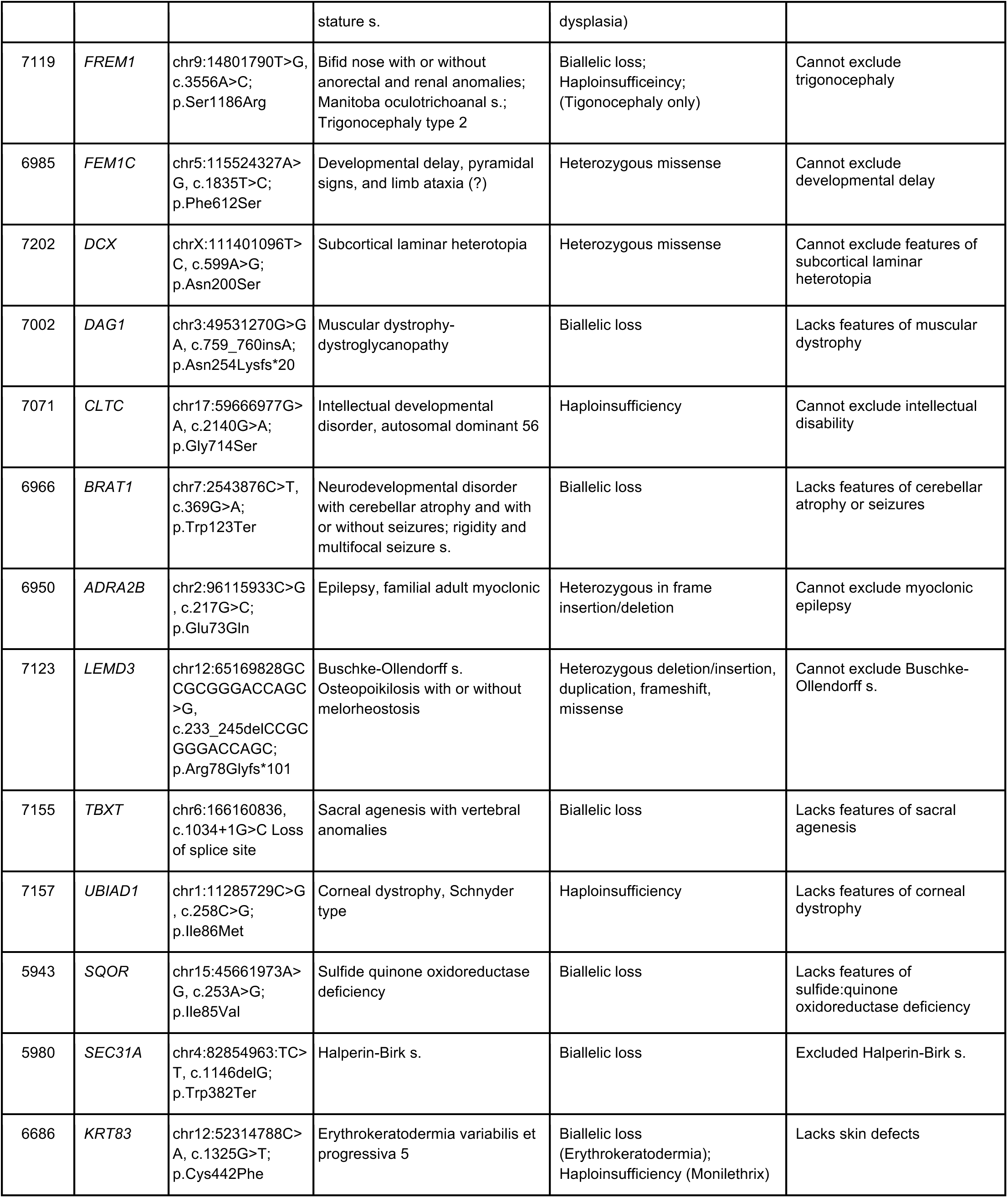
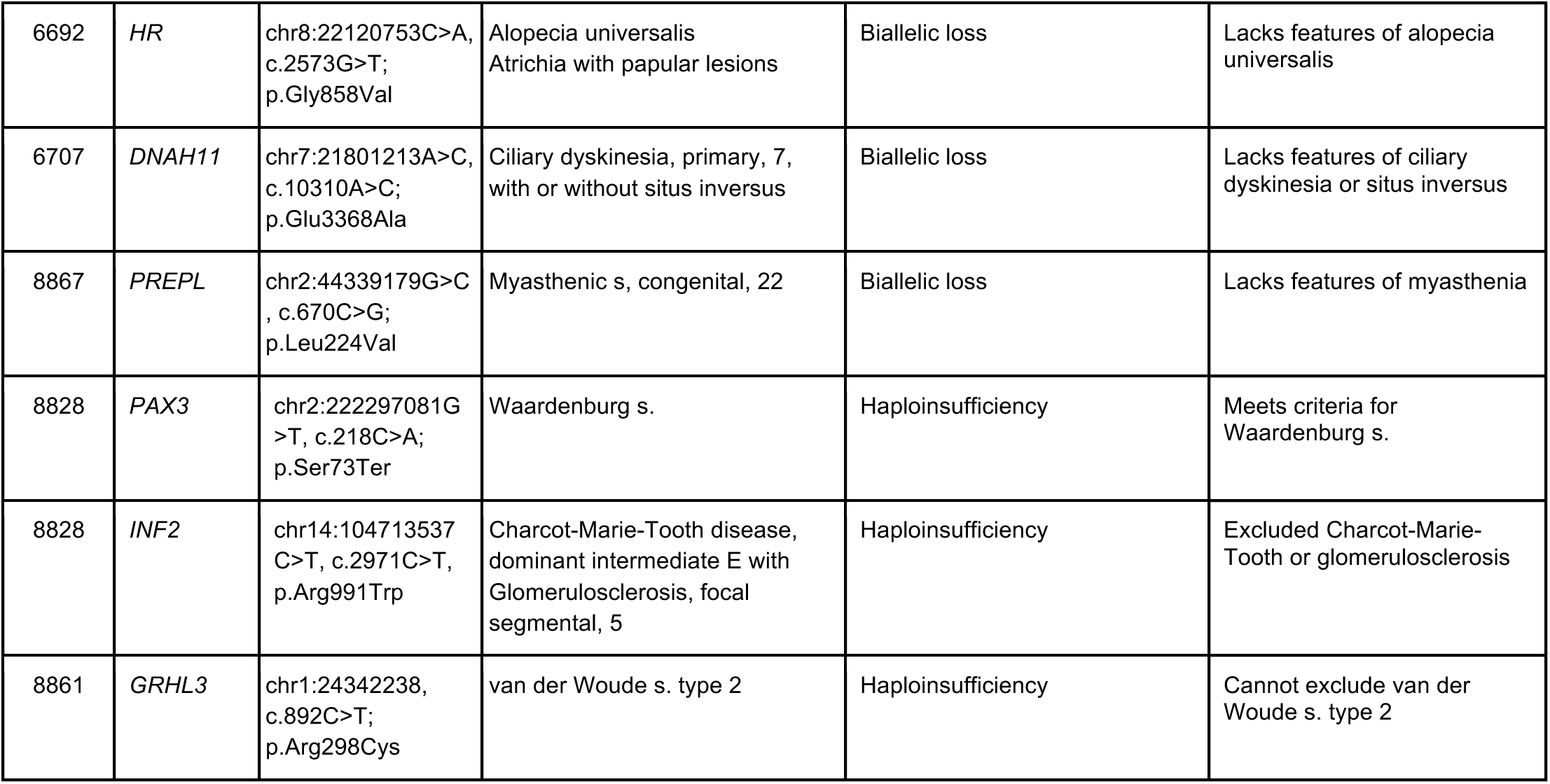
Phenotypic expansion of MM genes with DNM displaying syndrome and/or non syndrome disease features other than MM. Column 1: Family number, Column 2: mutated gene, Column3: Mutation identified for that gene and patient in our cohort, Column 4: Relevant OMIM entry for each gene. Column 5: type of mutation reported in OMIM leading to phenotype. Column 6: Evidence for or against concordance with the OMIM phenotype. Genomic coordinates are in hg38. s.: syndrome, question mark (?): association remains uncertain according to OMIM.

## Supplementary Notes

### Power calculations for effect size

To determine the adequate cohort size for identification of recurrently mutated genes in MM, we developed a computational model and power calculations to estimate effect size^77,78^. We first determined that with a sample size of 350 trios, observing such recurrence in two families would be significant genome-wide, with *P* value = 0.003, and from this calculated the false-discovery rate (FDR), assuming a baseline DNM rate among the ∼20,000 human genes (**Extended Data Fig. 1** gray dashed line). Then, leveraging data from a small NTD WES study in which DNM LOF mutations were identified in 20% of subjects, 34, and estimating that about 0.4% (i.e. 80/20,000) of gene knockouts in mouse show a MM-like phenotype, we estimate that there are 50–100 MM disease genes (*k*) to be discovered. Borrowing from previous literature ^13,21,79^, we anticipate that LOF variants will be enriched in patients vs. controls with a v ratio between 1.5 and 2.5. We ran 10,000 iterations to estimate the number of genes we will discover for various cohort sizes ranging from 100–1000 trios (**Extended Data Fig. 1**). For instance, with a cohort size of 400, if there are 50 genes to discover (i.e. *k* = 50) and with 2.5 times as many LOF variants in patients vs. control (i.e. v = 2.5), we expect ∼16 recurrently mutated genes to emerge. For the same cohort, if *k* = 100 and v = 1.5, we expect to discover ∼2 recurrently mutated genes. Given the recurrence found in the cohort of 43 trios ^15^, and our preliminary data, we anticipated our results will fall between these two scenarios. Notably, this model predicts that with 800 MM trios sequenced as we propose here, we expect to discover ∼ 5–10 recurrently mutated MM genes.

### Reproducible prediction of D-Mis-HC

To validate the high confidence damaging missense variant (D-Mis-HC) that was originally called by MetaSVM, we also applied MetaLR and REVEL to the DNMs. The D-Mis-HC was highly reproducible with the two additional annotation tools with recommended thresholds, MetaLR score > 0.5 and REVEL > 0.75 (**Extended Data Fig. 16c**-16d).

### Parents’ age of conception comparison

To confirm if the excessive DNM burden in MM compared to controls are independent from the ages of parents at the time of conception, we gathered dates of births from probands, fathers, and mothers. Trios that were able to track all information from three family members (MM: 79, control: 683) were collected, to compare the ages of conception of father and mother, with Wilcoxon rank-sum test. (two-sided). We could clearly confirm that both the ages of mothers and fathers in the MM cohort were significantly lower than those of controls (**Extended Data Fig. 2a**). Although the rate of de novo variants is known to be highly correlated with the age of parents, the excessive burden observed in our study is not derived from the difference of fathers’ and mothers’ age distribution.

### WGS evaluation for DNMs contributing to MM risk

From 101 additional trios and 1 quartet, we observed comparable de novo SNV and Indel rates with WGS data (**Extended Data Table 1**). Among them, 13 LGD and 39 D-Mis coding DNMs were observed. Also, 9 splicing disrupting variants (SpliceAI maximum delta score larger than 0.9) and 41 possible pathogenic noncoding DNMs (GREEN-VARAN level 4) were collected. However, there was no recurrent gene mutated within the WGS cohorts (see **Supplementary Data** for the full list of DNMs in WGS). Instead, three overlapped with damaging (LGD+D-Mis) DNMs with WES data were revealed: a stop-gain in KDM1A, a splice donor gain in ITPR3, and a variant in GRHL2 enhancer. We could find four de novo CNVs (> 100 kb) in WGS trios, four de novo deletions and duplications were observed (**Extended Data Table 2**). The large CNVs were confirmed with the parents’ sequence data (**Extended Data Fig. 3b**) and the duplicated or deleted regions were not recurrent. No de novo translocation or inversion was observed.

### Multiplexed Error Robust In Situ Hybridization (MERFISH) spatial transcriptomic expression pattern of a subset of DNM genes

We conducted Multiplexed Error Robust Fluorescent In Situ Hybridization (MERFISH) on the Vizgen platform using the methodology as described in Methods^64^. We segmented a total of 352,729 cells with uniform transcript density using cell segmentation. A subset of cells was excluded due to outlier cell volume (*n* = 9,922) or transcript density (*n* = 21,319), with the remaining cells divided into 63 distinct clusters based on gene expression using the Leiden algorithm, stratifying into 7 major cell types (Neuron, Neural crest, Neural Progenitor, Dorsal Root Ganglia, Pre epithelial to mesenchymal transition neural Crest progenitor, Mesoderm and Blood) in mouse E9.5 parasagittal sections. Cell types were defined by expression of marker genes selected based on previously published single cell transcriptomic studies^65–67^, employing ScType^80^ for unbiased cell typing from marker gene expression. Approximately 14% cells were flagged as ‘indeterminate’ type that could not be clearly categorized, likely due either to a state of transitioning between cell types or lacking expression of marker genes. We selected 36 DNM genes identified in MM subjects for multiplex spatial co-expression analysis (**Extended Data Fig. 4)**.

Percentage of gene expression vs cell type revealed that most of the candidate DNM genes show expression in multiple cell types during neural tube closure. There were some genes like *Rnd2* and *Add2* that were expressed predominantly in neural cells, *Celsr1* in neural progenitor, *Stab1* in pre-EMT-neural precursor cells, *Opalin* expressed in predominantly in blood (**Extended Data Fig. 5)**.

### Additional evidence for role of functionally implicated DNM genes in NTDs

The DNM genes in which the DNM mutation impairs protein function showed additional prior evidence that may suggest a role in human NTDs. TIAM1, which promotes actin assembly via the Arp2/3 complex^81^, is essential for neural tube closure in mice^82^, but not previously implicated in human NTD. TNK2, also known as ACK1 is a kinase associated with CDC42 that controls cellular protrusions associated with spinal neurulation in mouse^32^. VWA8 is a mitochondrial matrix-targeting ATPase, and notably mitochondrial dysfunction is implicated in both genetic and environmental murine models of NTD^49,83^.

### Clinical phenotypes of patients compared to OMIM

We assessed possible connections between DNM genes and associated OMIM phenotypes by recontacting subjects to inquire about clinical features listed in OMIM but possibly not described at the time of enrollment into our study. Resources used were Online Mendelian Inheritance in Man (OMIM), ClinVar, and PubMed. Out of the 192 DNMs found in our cohort 82 were found to have other OMIM phenotypes (43.8%). Genes found to have other OMIM phenotypes in individual families/subjects were further reviewed regarding the type of mutation compared with those reported in the clinical literature and compared with clinical status (**Supplementary Table 1**). Information available at the time of enrollment was used for clinical correlation along with follow up with the families via telephone or zoom to specifically inquire about features related to the published OMIM phenotypes. For some subjects like 6695, the de novo mutation occurred in a gene showing only recessive inheritance (*TIMM50*), so it was not surprising that the MM subject carrying a DNM in the gene did not report features of the OMIM entry. But for other subjects like 6313, the DNM occurred in a gene with the same or similar zygosity, and the patient showed features that cannot be distinguished from the reported OMIM entry, in this case Coffin-Siris features (i.e. developmental disability, abnormalities of the fifth (pinky) fingers or toes, and characteristic facial features), suggesting MM may be a phenotypic expansion of this syndrome.

### Estimation of number of genes in DNM contributing to MM risk

The estimation was based on previous publication^14^. Briefly, the total number of risk-associated genes (*C*) is calculated with the number of observed risk-associated mutations (*d* = 192) and recurrent mutations (*r* = 5, *PAX3*, *IRS1*, *ZSWIM6*, *BRSK2*, and *VWA8*).

Since 28.26% of the damaging DNMs (LGD + D-Mis) are contributing to the MM risk, *d* and *r* were normalized as 54.26 and 1.41. The number of observed risk-associated genes is *c* and *c_1_* refers to the number of genes mutated once and the probability of a newly added mutation hitting a previously mutated gene (*μ*) could be calculated as follows.

Considering the variation in effect size of individual DNMs are assumed to be 1 to minimize underestimation of set size, *C* could be estimated as 2,021 genes.

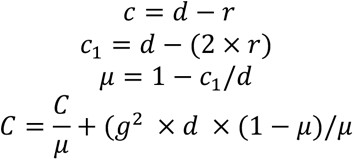

